# Impairments in goal-directed action and reversal learning in a proportion of individuals with psychosis: evidence for differential phenotypes in early and persistent psychosis

**DOI:** 10.1101/2021.08.31.21262937

**Authors:** Shuichi Suetani, Andrea Baker, Kelly Garner, Peter Cosgrove, Matilda Mackay-Sim, Dan Siskind, Graham K Murray, James G Scott, James P Kesby

**Author notes:** **Correspondence:** Dr. James P Kesby, Queensland Brain Institute, The University of Queensland, Brisbane, QLD 4072, Australia., (J.P.K). **Competing interests:** The authors have no competing interests.

## Abstract

Cognitive impairments in psychosis are one of the strongest predictors of functional decline. Cortico-striatal dysfunction may contribute to both psychosis and cognitive impairment in psychotic illnesses. The decision-making processes underlying goal-directed action and serial reversal learning can be measured and are sensitive to changes reflecting cortico-striatal dysfunction. As such, changes in decision-making performance may assist with predicting functional decline in people with psychosis. We assessed decision-making processes in healthy controls (N=34), and those with early psychosis (N=15) and persistent psychosis (N=45). We subclassified subjects based on intact/impaired goal-directed action. Compared with healthy controls (<20%), a large proportion (58%) of those with persistent psychosis displayed impaired goal-directed action, predicting poor serial reversal learning performance. Computational approaches indicated that those with persistent psychosis were less deterministic in their decision-making. Those with impaired goal-directed action had a decreased capacity to rapidly update their prior beliefs in the face of changing contingencies. In contrast, the early psychosis group included a lower proportion of individuals with impaired goal-directed action (20%) and displayed a different cognitive phenotype from those with persistent psychosis. These findings suggest prominent decision-making deficits, indicative of cortico-striatal dysfunction, are present in a large proportion of people with persistent psychosis while those with early psychosis have relatively intact decision-making processes compared to healthy controls. It is unclear if there is a progressive decline in decision-making processes in some individuals with psychosis or if the presence of decision-making processes in early psychosis is predictive of a persistent trajectory of illness.

## 1. INTRODUCTION

People with psychosis suffer from a range of cognitive impairments, including deficits in working memory, verbal/visual learning, reasoning and problem solving (Marder, 2006). Cognitive impairments are strong predictors of functional outcomes across psychiatric diagnoses (Crouse et al., 2020), particularly for those with psychosis (Hochberger et al., 2020). Therefore, there is a strong interest in identifying cognitive features that help to predict illness trajectories for these individuals (Nelson et al., 2017; Reichenberg et al., 2010).

Psychotic disorders are heterogeneous in both neurobiology and symptom profile. However, robust evidence highlights a strong link between psychosis and subcortical dopamine systems (Kesby et al., 2018; Li et al., 2020). Positron emission tomography (PET) studies have demonstrated that excessive dopamine signaling in the associative striatum underlies psychosis, and potentially cognitive deficits (Conn et al., 2020; Ersche et al., 2011; McCutcheon et al., 2018). Imaging protocols suggest that functional associative striatal abnormalities may serve as a biomarker for psychotic disorders (Li et al., 2020). The associative striatum receives a rich set of connections from higher-order cortical regions and selectively gates incoming cortical information (Conn et al., 2020). This enables associative striatal networks to modulate information flow in order to generate and adapt responses for action selection (i.e., decision-making) (Sharpe et al., 2018). Decision-making problems are common in those with psychotic disorders (Adida et al., 2011; Bates et al., 2002; Chudasama and Robbins, 2006; Morris et al., 2018; Pantelis et al., 2004) and may be due to dysfunctional cortico-striatal circuits. After dopamine stimulation (Clatworthy et al., 2009) and in psychosis (Dandash et al., 2014; Morris et al., 2015; Sarpal et al., 2015), changes in the functional connectivity and activation of the associative striatum are evident suggesting a causative role in decision-making deficits. Therefore, there may be common neurobiological mechanisms underlying psychosis and decision-making impairments.

Decision-making involves the contribution of a range of brain areas and circuits. We have proposed that two tests, outcome-specific devaluation and serial reversal learning, represent a behavioral approach sensitive to associative striatal dysfunction (Conn et al., 2020; Kesby et al., 2018). Altered activation of the associative striatum (caudate) underlies impairments in outcome devaluation in people with schizophrenia (Morris et al., 2015). In reversal learning studies, increases in the associative striatal dopamine levels of healthy individuals have been shown to correlate with decreased performance (Clatworthy et al., 2009). Deficits in reversal learning have also been observed in those with first episode psychosis (Murray et al., 2008), and have been associated with thought disorder in schizophrenia (Pantelis et al., 2004).

Whether deficits in one task predict deficits in the other has not been investigated. Such findings would provide evidence that associative striatal dysfunction may contribute to the cognitive deficits observed in those with psychosis. The aims of the present study were (i) to establish whether people with persistent psychosis display deficits in both outcome devaluation and reversal learning compared with healthy individuals, and (ii) to assess behavioral performance in those with early psychosis to determine if decision-making deficits are evident early in the illness course.

## 2. MATERIALS AND METHODS

### 2.1 Participants

A total of 94 participants, between 18-50 years of age, were classified into three groups based on psychiatric history. Healthy controls had no diagnosis of a psychotic disorder and had not experienced a psychotic episode (N=34). Those diagnosed with a psychotic disorder were separated into two groups: early psychosis (within 6 months of receiving initial antipsychotic treatment; N=15) and persistent psychosis (those not fitting the criteria for early psychosis; N=45). See **Table 1** for general psychiatric characteristics for those with psychosis (see *Table S1* for Positive and Negative Syndrome Scale [PANSS] sub scores). All diagnoses were based on clinician descriptions in the patient’s health records. Therefore, patients with ambiguous records (i.e., “unspecified non-organic psychosis”) are listed as “Other” rather than nonaffective or affective diagnoses. Detailed inclusion criteria are described in the *Supplementary Methods*.

**Table 1.**
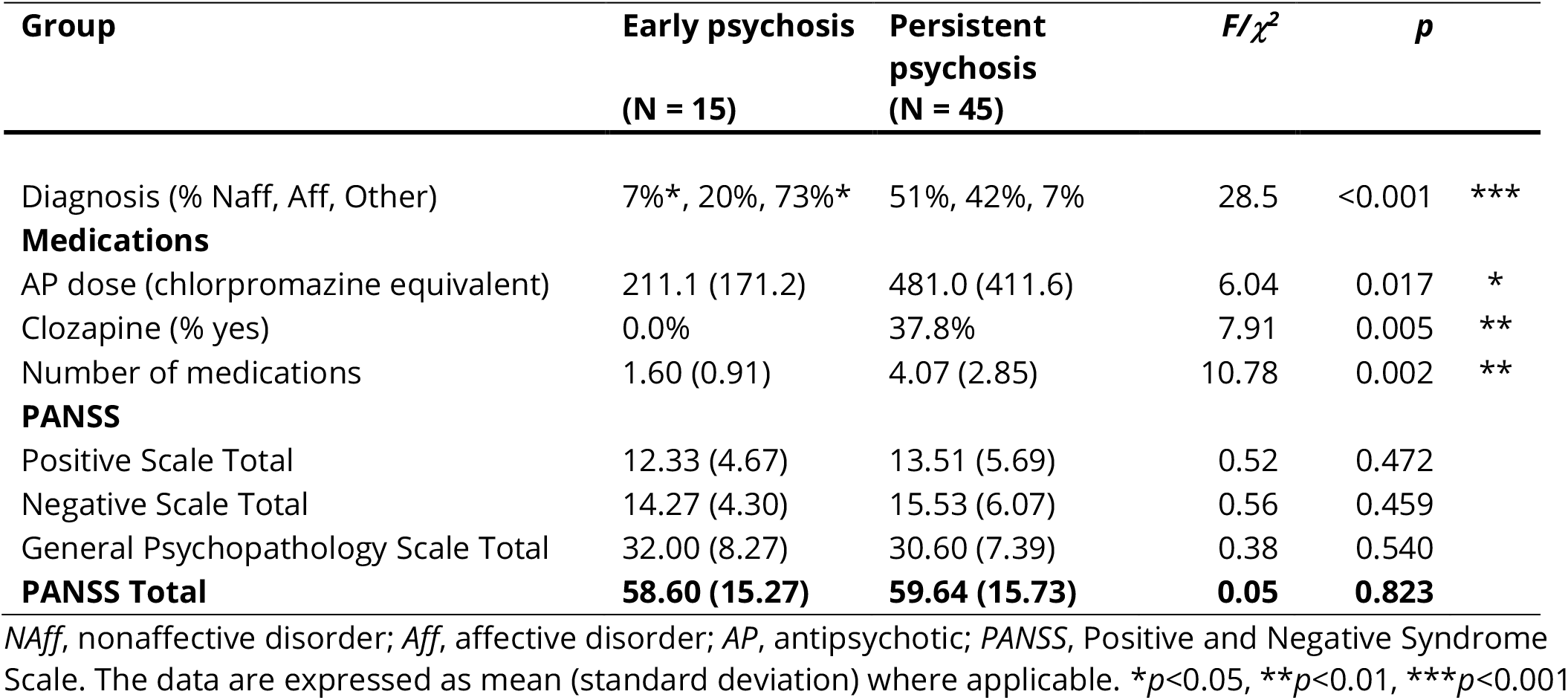
Psychiatric characteristics and symptom assessments

| Group | Early psychosis<br>(N = 15) | Persistent<br>psychosis<br>(N = 45) | $F/\chi^2$ | $p$ | |
| --- | --- | --- | --- | --- | --- |
| Diagnosis (% Naff, Aff, Other) | 7%*, 20%, 73%* | 51%, 42%, 7% | 28.5 | <0.001 | *** |
| **Medications** |  |  |  |  |  |
| AP dose (chlorpromazine equivalent) | 211.1 (171.2) | 481.0 (411.6) | 6.04 | 0.017 | * |
| Clozapine (% yes) | 0.0% | 37.8% | 7.91 | 0.005 | ** |
| Number of medications | 1.60 (0.91) | 4.07 (2.85) | 10.78 | 0.002 | ** |
| **PANSS** |  |  |  |  |  |
| Positive Scale Total | 12.33 (4.67) | 13.51 (5.69) | 0.52 | 0.472 |  |
| Negative Scale Total | 14.27 (4.30) | 15.53 (6.07) | 0.56 | 0.459 |  |
| General Psychopathology Scale Total | 32.00 (8.27) | 30.60 (7.39) | 0.38 | 0.540 |  |
| **PANSS Total** | **58.60 (15.27)** | **59.64 (15.73)** | **0.05** | **0.823** |  |

*NAff*, nonaffective disorder; *Aff*, affective disorder; *AP*, antipsychotic; *PANSS*, Positive and Negative Syndrome Scale. The data are expressed as mean (standard deviation) where applicable. \* $p < 0.05$ , \*\* $p < 0.01$ , \*\*\* $p < 0.001$

### 2.2 Procedures and experimental design

All procedures were approved by the Royal Brisbane and Women’s Hospital, and University of Queensland Human Research Ethics Committees (HREC/17/QRBW/168). Participants were remunerated $40AUD (see *Supplementary methods*). Premorbid and current IQ was assessed using the Test of Premorbid Functioning (TOPF; Pearson Clinical, Sydney, Australia) and Wechsler Abbreviated Scale of Intelligence, second edition (WASI-II; Pearson Clinical). Substance use was assessed using a Substance Misuse Scale (Duhig et al., 2015). The cognitive tasks were run using PsychoPy v3 (Peirce et al., 2019) with stimuli being displayed on a computer monitor. Responses were recorded on a joystick box (Fighting stick mini 4; Hori Co. Ltd, Yokohama, Japan).

### 2.3 Outcome-specific devaluation task

We used an outcome devaluation task (**Figure 1**) adapted from Morris et al. (2015).

**Figure 1.**
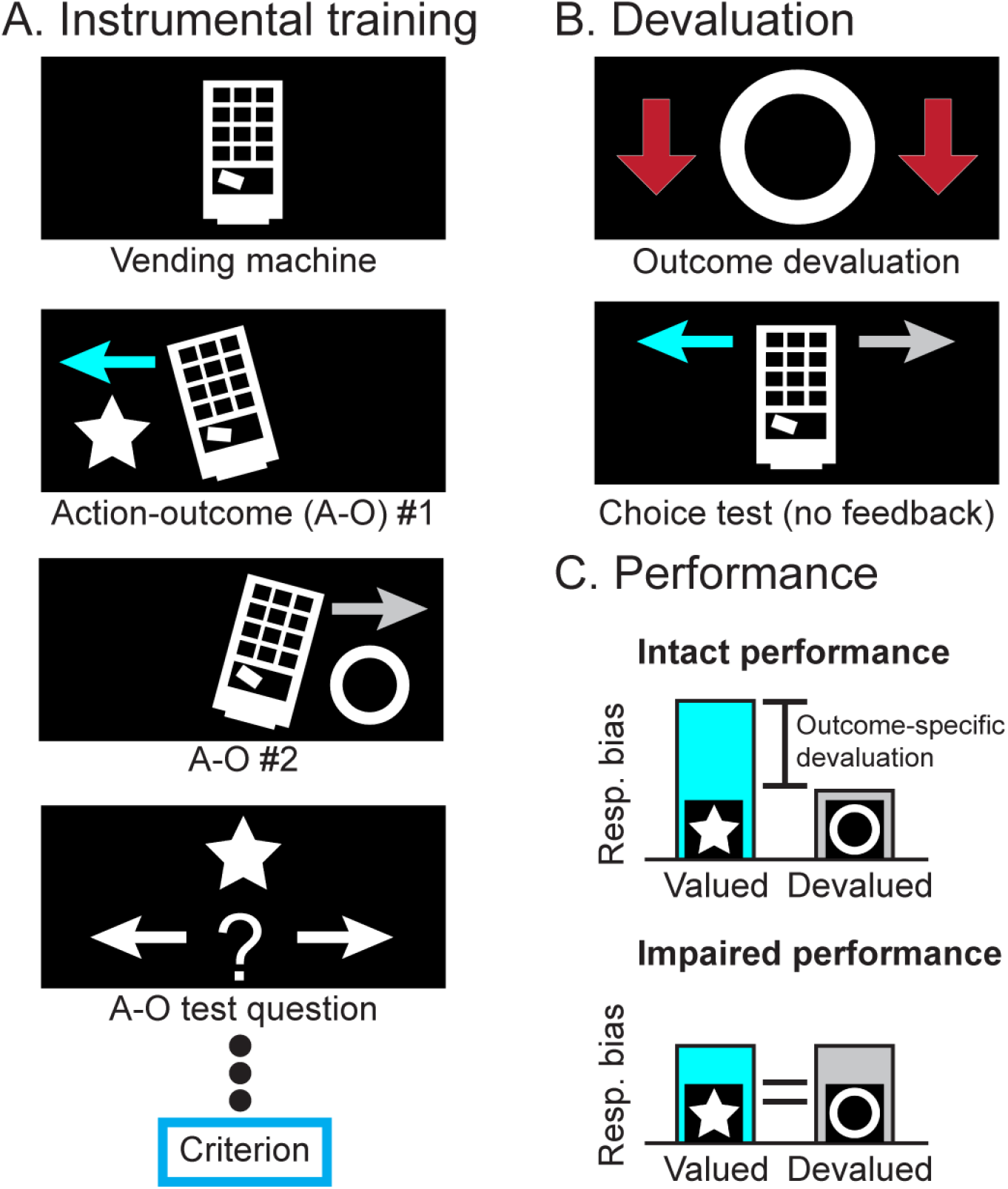
Outcome-specific devaluation task. Participants are trained to learn two action-outcome (A-O) associations by tilting a vending machine left or right with a joystick (**A**). To confirm they have learned these relationships, a test question is presented after three stimuli presentations. Participants must correctly answer 6 consecutive test questions to reach criterion. Following instrumental training, participants are informed that one outcome is now worth less credits (outcome devaluation) and undergo a choice test (**B**). In the choice test the participant can tilt the vending machine left or right but receives no feedback. Performance is assessed using the responses (resp.) and response bias between the valued and devalued actions (**C**). A significant bias in responding towards the valued outcome (intact performance) indicates intact goal-directed action, whereas a lack of this bias indicates impaired performance.

#### 2.3.1 Instrumental training

Participants were told that three tokens (visual stimuli) were of equal value. Using a 7-point Likert scale, participants rated each token (*Fig S1*) based on how valuable they considered them to be, and their motivation to earn tokens. Training involved liberating two of the tokens from a virtual vending machine. The joystick was moved left or right, with 5-10 consecutive responses (drawn randomly) in one direction required to earn the associated token (e.g., star or circle). After every three rounds, a question was posed to assess participants’ understanding of the association between action and reward. After getting six questions correct in a row, instrumental training ended.

#### 2.3.2 Devaluation test

Participants were informed that one of the tokens had been counterfeited (counterbalanced) and was therefore less valuable. Participants were instructed to tilt the vending machine in order to earn the associated tokens, and that their actions in this stage would dictate their monetary compensation. The virtual machine was displayed for 10 blocks (12 seconds) and could be tilted at will during each block. Aside from visually tilting the vending machine, no feedback was presented. Subsequently, participants were asked probe questions about which outcome was associated with which action and rerated the value of each token and their motivation to receive tokens.

### 2.4 Serial reversal learning task

For the reversal learning task (**Figure 2**), all stimulus pairs were binary images (see *Fig S2*) and all combinations counterbalanced (for detailed methodology and training stages see *Supplementary methods*).

**Figure 2.**
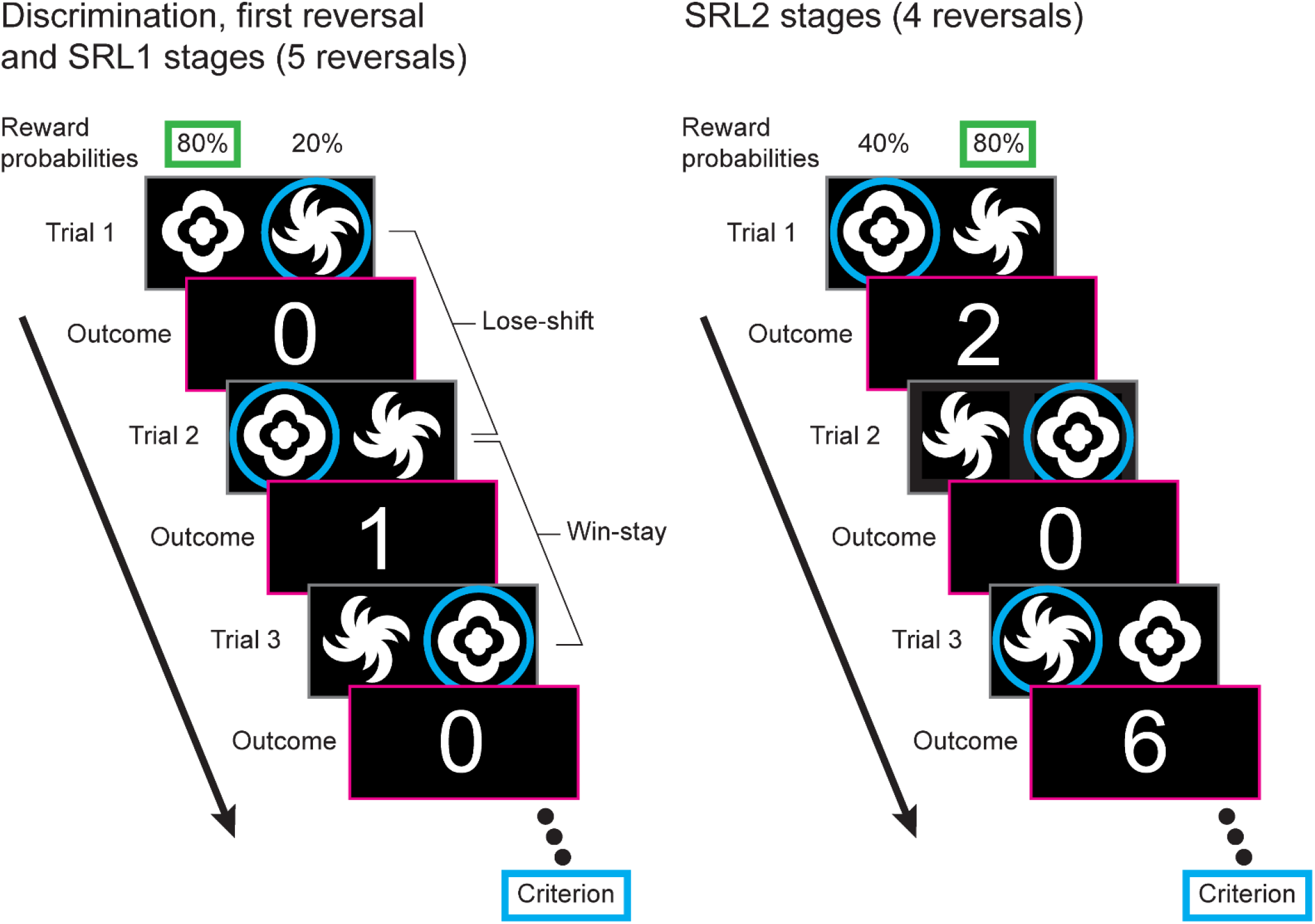
Serial reversal learning task. For each trial participants are presented with two stimuli (which remain the same throughout the entire task). For the initial discrimination, first reversal and serial reversal learning (SRL) 1 stages (left), these are probabilistically rewarded with one stimulus rewarded on 80% of trials (cloud-like stimuli in this example) and the other at 20% (spiral-like stimuli in this example). After selecting a stimulus (blue circle) the credit reward is displayed onscreen (0 or 1 credit). A Lose-shift is recorded when the participant changes the alternative stimulus after a loss (i.e., after a loss on Trial 1, 0 credits, the other stimulus is selected for Trial 2), and a Win-stay is recorded when the participant selects the same stimulus after a win (i.e., Trial 2 wins 1 credit, and the same stimulus is selected for Trial 3). Criterion is reached when 6 consecutive choices of the higher probability stimulus are made, and then, for the next stage, the stimulus probabilities are reversed. For the SRL2 stages (right), the probability of receiving a reward on the poorer stimulus are increased to 40% and participants can receive either 2 or 6 credits for a win (equal probability).

#### 2.4.1 Probabilistic reversal learning

Participants underwent a probabilistic reversal learning task consisting of 11 stages; initial discrimination (1 stage), initial reversal (1 stage), and serial reversal learning phase 1 (SRL1; 5 stages) and phase 2 (SRL2; 4 stages). Each featured the same pair of stimuli but varying in reward rate (probabilistic) and outcome (credits). For the first seven stages, the probabilistic reward contingencies were set at 80/20, meaning that the target stimulus was rewarded 80% of the time, and the non-target stimulus was rewarded 20% of the time. One credit was earned for a rewarded trial and 0 credits for a non-rewarded trial. For the SRL2 stages, the contingencies were set at 80/40, increasing the task difficulty by providing more misleading feedback. Two or six credits were earned for a rewarded trial (equal probability) and 0 credits for a non-rewarded trial. Criterion for progressing from each stage was 6 correct responses in a row.

#### 2.4.2 Reversal learning performance measures and strategies

General performance measures included total trials to criterion, perseveration (number of errors in the first 6 trials after a reversal), and response rates. Whether a subject selected the same stimulus after attaining a reward (Win-stay) or selected the alternative stimulus after a loss (Lose-shift) was quantified as a proportion of the total applicable trials.

### 2.5 Computational modeling and simulation

The underlying cognitive processes in reversal learning were calculated by modeling latent task variables using the hBayesDM package for R (version 3.6 [Platform: x86_64-w64-mingw32/x64 (64-bit)] on Windows 10 v1809) developed by Ahn et al. (2017). Two models that had previously shown a good fit to reversal learning behavior were examined (den Ouden et al., 2013): an experience-weighted attraction model (EWA) and a reward/punishment learning model (RP). Parameters included learning rate (EWA *phi*), experience decay (EWA *rho*; how quickly prior information is updated), reward learning rate (RP), punishment learning rate (RP), and inverse temperature (EWA and RP; the deterministic or exploratory nature of the choices made). The RP learning parameters were inverted (e.g., 1-parameter value) to maintain consistency with EWA learning rates. The estimated inverse temperature parameters using both models showed a high correlation (*Fig S3*), demonstrating that the models have good concordance.

To establish which parameters were responsible for alterations in performance, we simulated performance after the manipulation of each individual parameter in order to generate ‘hypothetical’ outcomes based on the model-driven performance (N=20/group).

### 2.6 Data analysis

Binary variables were examined using χ2-tests and continuous variables used analysis of variance (ANOVA) with *Group* as the independent variable (with repeated measures where necessary). The discrimination ratio and response rates for outcome devaluation were also analyzed using within-group paired *t*-tests to confirm significant goal-directed action. To classify subjects as having intact or impaired goal-directed action we performed hierarchical clustering analyses using Ward’s method and Squared Euclidean distance. Variables (preference ratio and response rates for the valued action on outcome devaluation) were transformed using Z scores. Computational simulations were compared using ANOVAs and then to control simulations using the Dunnett’s test for multiple comparisons. All statistical analyses were performed with IBM SPSS Statistics 26 (Armonk, NY, USA). When appropriate, post hoc comparisons were performed using Šídák corrections. Results are expressed as mean ± standard error of the mean (SEM). Differences were considered statistically significant at *p*<0.05. Preference and response bias figures were made with code adapted from (van Langen, 2020).

## 3. RESULTS

### 3.1 Decision-making in those with early psychosis

We selected a subgroup of 15 age- and sex-matched controls to compare with early psychosis subjects. **Table 2** provides the demographics, IQ and substance use in these groups (see *Table S2* for details).

**Table 2.**
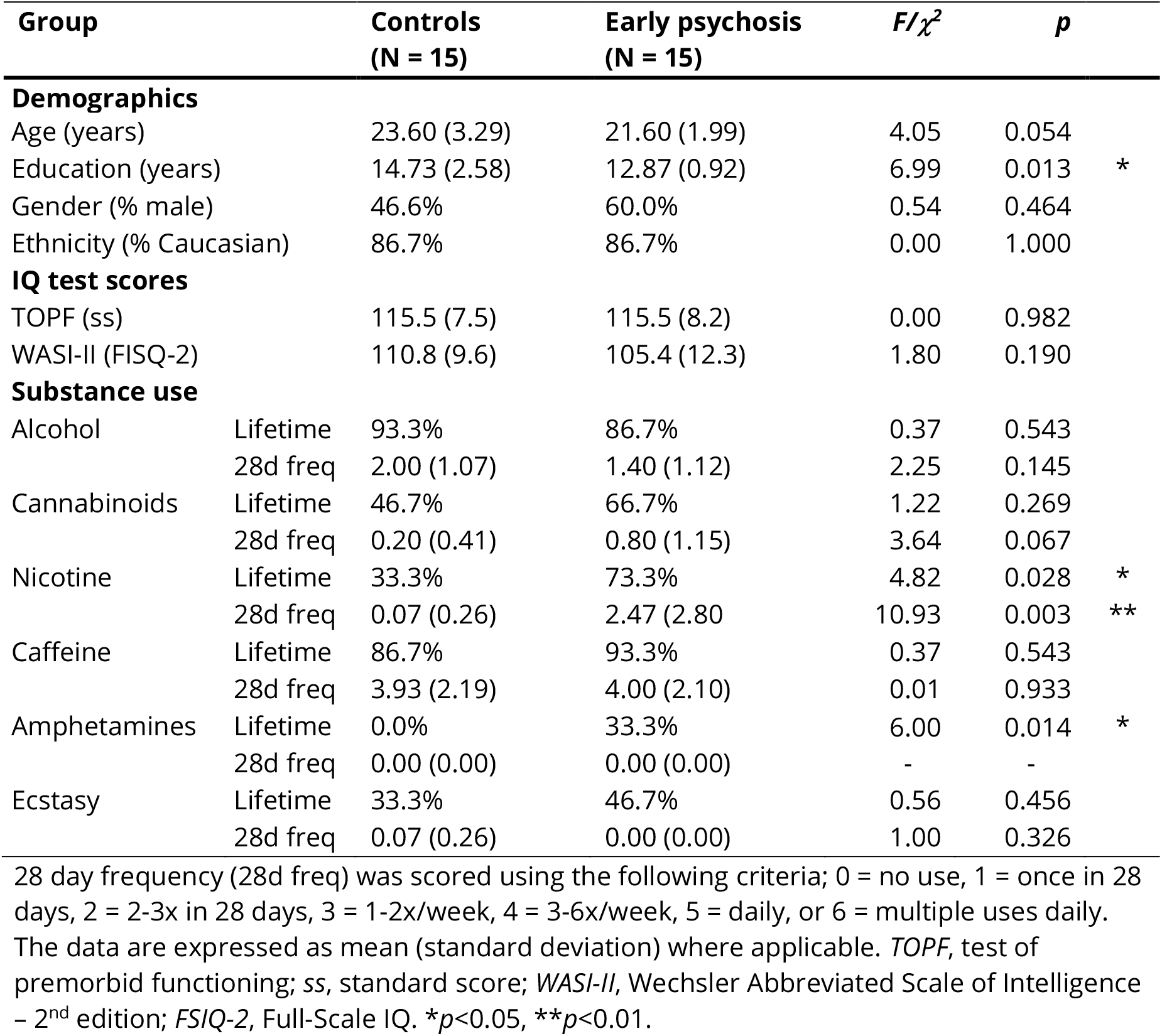
Demographics, IQ and substance use characteristics for age-matched control subjects and those with early psychosis.

#### 3.1.1 Goal-directed action is intact in those with early psychosis

Both young control and early psychosis groups showed a significant bias in preference towards the valued response (**Figure 3A**; for all comparisons see *Table S3*). There were no significant differences between groups in the response rates (**Figure 3B**). Both groups showed a significant decrease in their rating for the devalued versus the valued outcomes after devaluation (**Figure 3C**; young controls, *t*_14_ = 4.5, *p* <0.001; early psychosis, *t*_14_ = 3.7, *p* <0.01).

**Figure 3.**
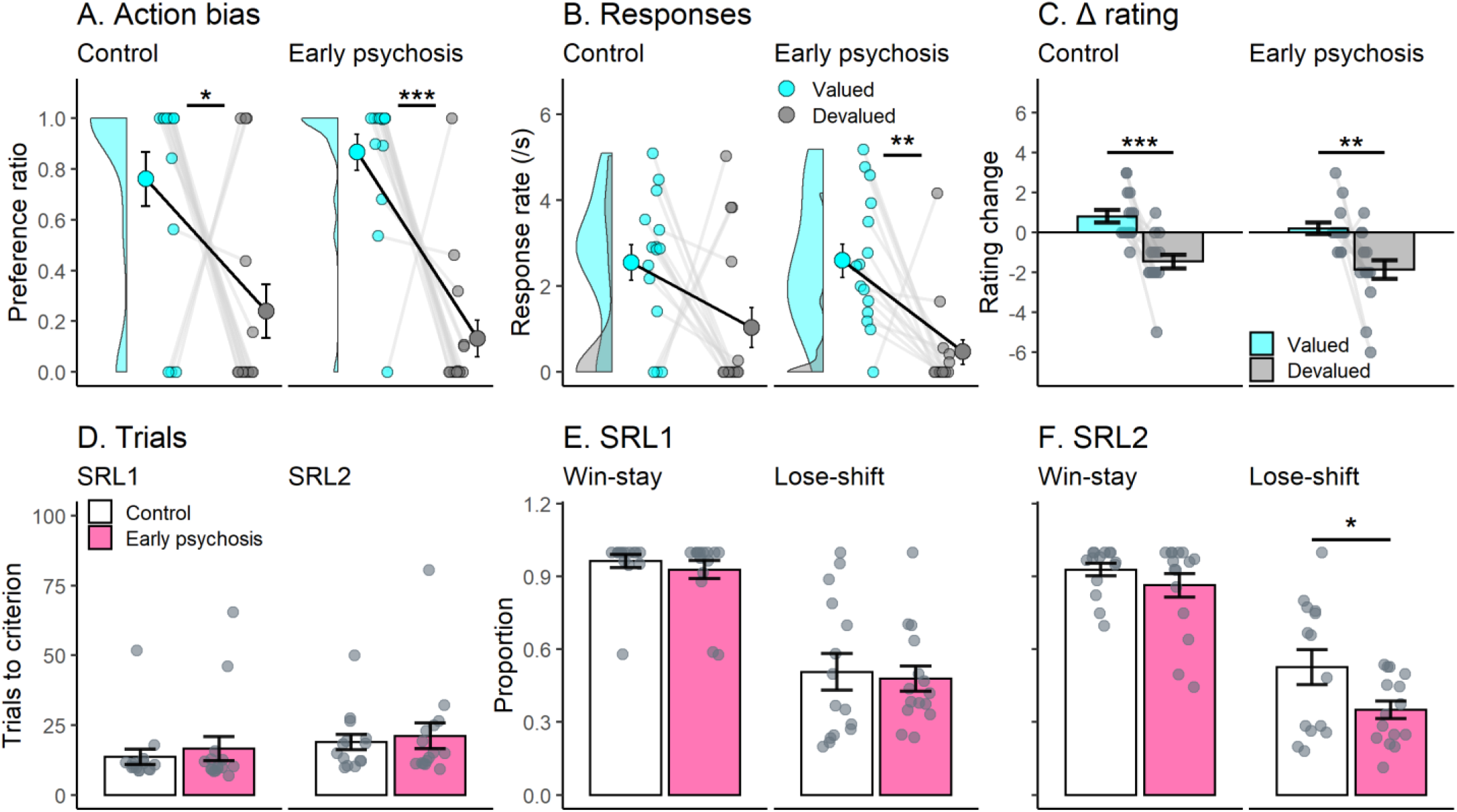
Decision-making in those with early psychosis. Comparison of performance in the outcome devaluation and reversal learning tasks in healthy age-matched controls and those with early psychosis. For outcome devaluation, early psychosis subjects showed a stronger bias (**A**) and response (**B**) towards the valued outcome after devaluation (**C**) than controls. For serial reversal learning, early psychosis subjects showed similar performance to controls when looking at trials to criterion for SRL1 and SRL2 stages (**D**). Strategy use in the SRL1 (**E**) and SRL2 (**F**) stages were similar to controls, but early psychosis subjects exhibited less use of Lose-shift strategies in the SRL2 stage suggesting they were less able to modify their decisions after negative feedback. Panels **A** and **B** feature frequency histograms (left of summary data) for the valued responses (blue, **A** and **B**) and devalued responses (grey, **B** only). Data are displayed as the mean ± standard error. \**p*<0.05, \*\**p*<0.01, \*\*\**p*<0.001.

#### 3.1.2 Reversal learning in those with early psychosis reveals potential changes in punishment learning

There were no significant differences observed between the control and early psychosis groups in the trials to criterion for any stage (**Figure 3D**; for all comparisons see *Table S4*). Strategies used during SRL1 were similar between controls and those with early psychosis (**Figure 3E**) but early psychosis subjects shifted less after losses (i.e., decreased Lose-shift use) during the SRL2 stage (**Figure 3F**; *F*_1,27_=4.8, *p*<0.05). Controls maintained a similar level of Lose-shift use between the SRL1 and SRL2 stages (*t*_13_ = 0.0, *p* =0.99), whereas early psychosis subjects significantly decreased their Lose-shift use in the SRL2 stage (*t*_13_ = 2.6, *p* <0.05). These data indicate that early psychosis subjects adapt differently to changing contingencies compared with controls.

### 3.2 Decision-making in those with persistent psychosis

Compared with controls, persistent psychosis subjects had significantly fewer years of education, and a lower average premorbid and current IQ (See **Table 3** for demographics, IQ and substance use). The persistent psychosis group also had a higher level of lifetime use for multiple substances (see *Table S5* for details).

**Table 3.**
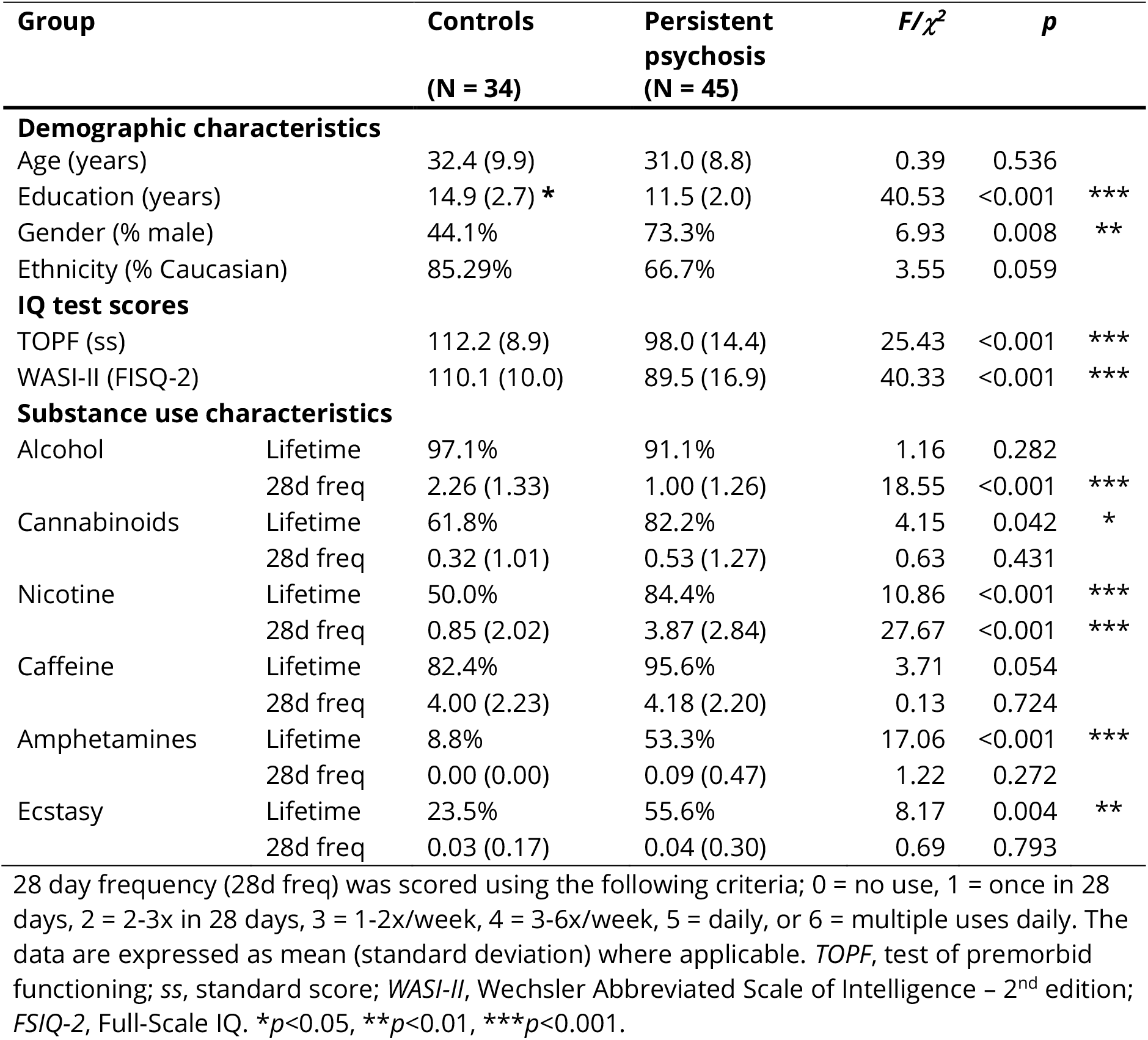
Demographics, IQ and substance use characteristics.

#### 3.2.2 Goal-directed action is impaired in a large proportion of those with persistent psychosis

Both control and persistent psychosis groups showed a significant bias in preference towards the valued response (**Figure 4A**) and in the rate of responding towards the valued response (**Figure 4B**). However, both the preference (*F*_2,78_=6.9, *p*<0.05), and rate of responding (*F*_2,78_=12.8, *p*<0.001), for the valued response was significantly lower in persistent psychosis subjects compared with controls (for all comparisons see *Table S6*). These data confirm that persistent psychosis subjects have deficits in goal-directed action (Morris et al., 2018; Morris et al., 2015). Decreases in goal-directed action were not due to changes in reward valuation. Both groups showed a significant decrease in their rating for the devalued versus the valued outcomes after devaluation (**Figure 4C**; controls, *t*_33_ = 8.0, *p* <0.001; persistent psychosis, *t*_43_ = 5.6, *p* <0.001). There was no significant difference between groups in the average number of correct responses for the probe questions, indicating that both groups recollected the action-outcome associations.

**Figure 4.**
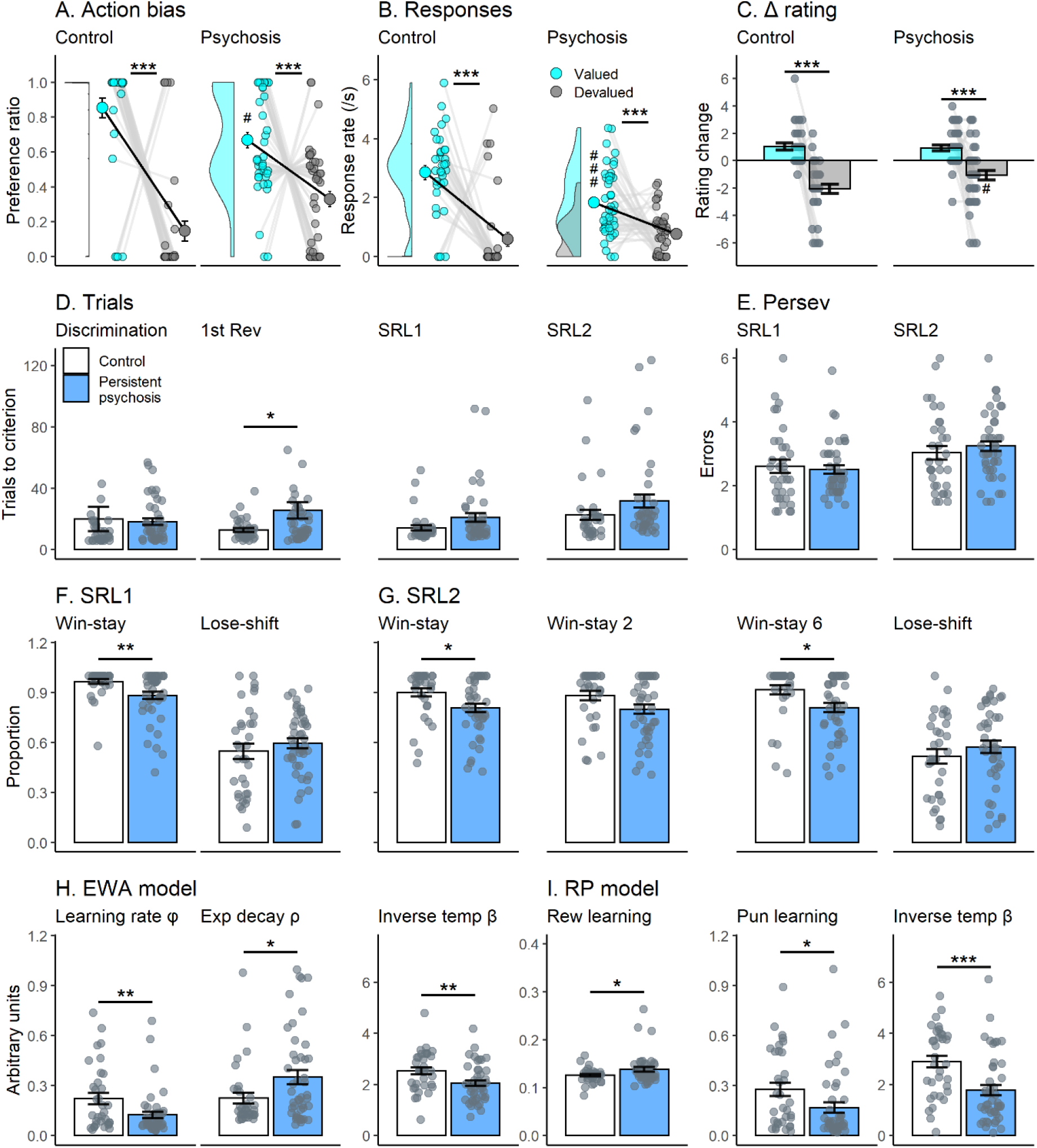
Decision-making performance in those with persistent psychosis. Comparison of outcome devaluation and reversal learning performance in healthy controls and those with persistent psychosis. For outcome devaluation, persistent psychosis subjects showed a significant bias (**A**) and response (**B**) towards the valued outcome after devaluation (**C**). However, the preference towards the valued outcome was significantly less than that observed in controls and featured a bimodal frequency distribution as seen in the frequency histogram along the Y Axis (**A**). For serial reversal learning, persistent psychosis subjects took significantly more trials to reach criterion (**D**) for the first reversal and trended towards the same for the SRL1 and SRL2 stages. These increases were not associated with changes in the number of perseverative errors (**E**). The strategies used for the SRL1 (**F**) and SRL2 (**G**) showed a similar pattern, with persistent psychosis subjects using fewer Win-stays than control subjects. Differences in computational modeling parameters were observed for all parameters in the EWA (**H**) and RP (**I**) models. *Note: RP reward and punishment learning are inverted to match the EWA learning rate (i.e*., *1 – parameter value)*. Data are displayed as the mean ± standard error. \**p*<0.05, \*\**p*<0.01, \*\*\**p*<0.001. ^#^*p*<0.05, ^###^*p*<0.001 compared with controls.

#### 3.2.3 Persistent psychosis subjects switch more after rewards in serial reversal learning

There were significant differences between groups for the trials to criterion in the first reversal (*F*_1,76_=4.3, *p*<0.05), and a trend in the SRL1 stage (*F*_1,76_=3.8, *p*=0.056), due to an increase in the average trials required for the persistent psychosis group compared with controls (**Figure 4D**; for all comparisons see *Table S7*). These were not accompanied by alterations in the number of perseverative errors (**Figure 4E**). The proportion of Win-stay, but not Lose-shift, use was significantly different between groups in the SRL1 stage (**Figure 4F**; *F*_1,76_=9.0, *p*<0.01) and the SRL2 stage (**Figure 4G**; *F*_1,69_=5.4, *p*<0.05). Those with persistent psychosis used Win-stay strategies less than controls, particularly after winning a 6 in the SRL2 stage (*F*_1,69_=6.2, *p*<0.05).

Computational modeling indicated that a range of decision-making processes were different between persistent psychosis subjects and controls. For the EWA model (**Figure 4H**), there were significant differences between groups for all parameters, with persistent psychosis subjects having a lower learning rate (*F*_1,76_=7.2, *p*<0.01), higher experience decay (*F*_1,76_=4.6, *p*<0.05) and lower inverse temperature (*F*_1,76_=8.6, *p*<0.01). Lower learning rate values indicate a bias toward using more recent information rather than past outcomes, with lower inverse temperature values reflecting less deterministic or more exploratory decision-making. Higher experience decay values indicate a slower decay or updating of experience weight with changing contingencies. For the RP model (**Figure 4I**), there were also significant differences between groups for all parameters, with persistent psychosis subjects having lower punishment learning (*F*_1,76_=5.0, *p*<0.05), higher reward learning (*F*_1,76_=4.7, *p*<0.05) and lower inverse temperature (*F*_1,76_=12.8, *p*<0.001). As with the EWA learning rate parameter, lower learning rates in the RP model reflect a bias toward using more recent information rather than past outcomes. Therefore, the observed decrease in the EWA learning rate parameter in those with persistent psychosis appears to be driven specifically by altered punishment learning.

### 3.3 A large proportion of those with persistent psychosis have broad decision-making deficits

Given the bimodal distribution for the valued lever preference in the persistent psychosis group (**Figure 4A**), we used hierarchical clustering analyses to classify each group into intact and impaired goal-directed action subgroups (see **Figure 1C**). The proportion of impaired subjects was greatest in the persistent psychosis group (controls, 28 intact/6 impaired; early psychosis, 12 intact/3 impaired; persistent psychosis, 18 intact/25 impaired). Low numbers prevented some comparisons (see *Tables S8-S10* for control/early psychosis intact/impaired data), therefore analyses included the control intact (n=25), and persistent psychosis intact (n=18) and impaired (n=25) subgroups. The demographical and psychiatric information for these groups can be found in **Tables 4 and 5** (see *Table S11* and *Table S12* for details).

**Table 4.**
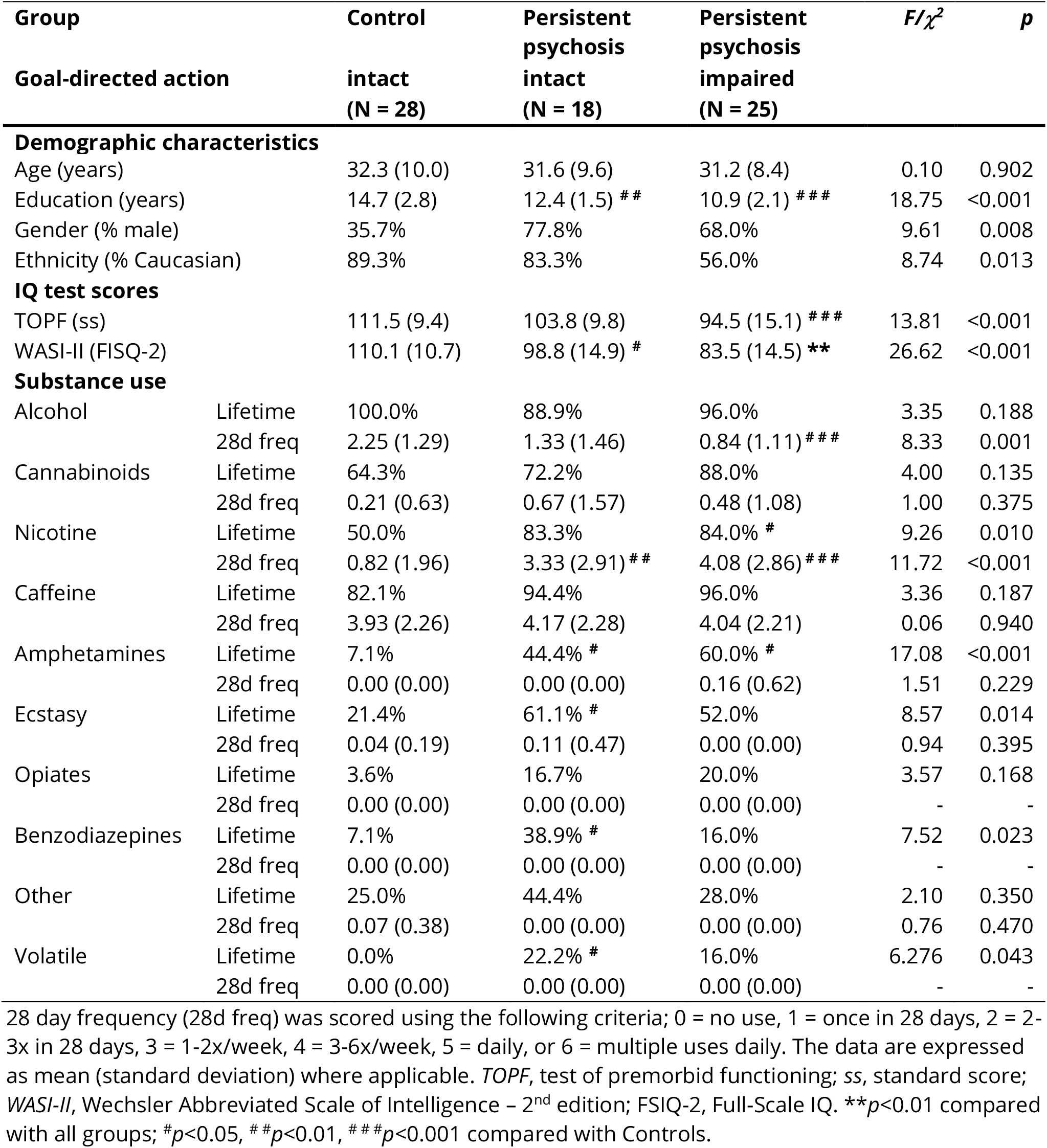
Demographics, IQ and substance use characteristics.

**Table 5.**
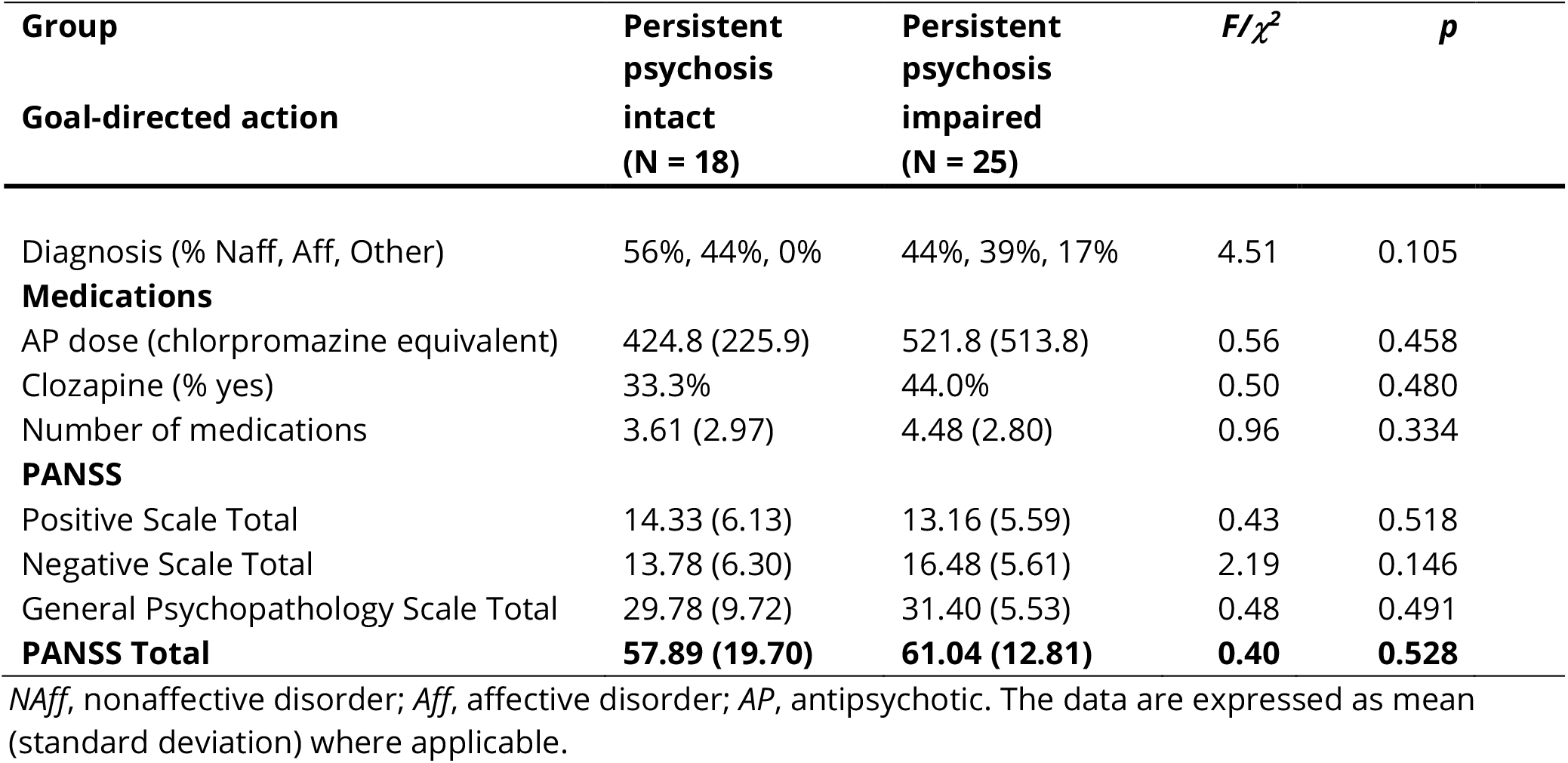
Psychiatric characteristics and symptom assessments of those with persistent psychosis split for intact or impaired goal-directed action

| **Group** | **Persistent psychosis intact (N = 18)** | **Persistent psychosis impaired (N = 25)** | **$F/\chi^2$** | **$p$** |
| --- | --- | --- | --- | --- |
| **Goal-directed action** |  |  |  |  |
| Diagnosis (% Naff, Aff, Other) | 56%, 44%, 0% | 44%, 39%, 17% | 4.51 | 0.105 |
| **Medications** |  |  |  |  |
| AP dose (chlorpromazine equivalent) | 424.8 (225.9) | 521.8 (513.8) | 0.56 | 0.458 |
| Clozapine (% yes) | 33.3% | 44.0% | 0.50 | 0.480 |
| Number of medications | 3.61 (2.97) | 4.48 (2.80) | 0.96 | 0.334 |
| **PANSS** |  |  |  |  |
| Positive Scale Total | 14.33 (6.13) | 13.16 (5.59) | 0.43 | 0.518 |
| Negative Scale Total | 13.78 (6.30) | 16.48 (5.61) | 2.19 | 0.146 |
| General Psychopathology Scale Total | 29.78 (9.72) | 31.40 (5.53) | 0.48 | 0.491 |
| **PANSS Total** | **57.89 (19.70)** | **61.04 (12.81)** | **0.40** | **0.528** |

*NAff*, nonaffective disorder; *Aff*, affective disorder; *AP*, antipsychotic. The data are expressed as mean (standard deviation) where applicable.

#### 3.3.1 Impaired goal-directed action is not due to impaired reward valuation

**Figures 5A** and **Figure 5B** show the preference and response rates for subgroups with intact and impaired goal-directed action (for all comparisons see *Table S13*). All groups showed a significant reduction in their rating for the devalued compared with valued outcomes after devaluation indicating that impairments in reward valuation do not underly impaired goal-directed action (**Figure 5C**). There was a significant difference between groups in the magnitude of change for the devalued outcome (*F*_2,68_=7.8, *p*<0.001). Persistent psychosis subjects with impaired goal-directed action had a smaller decrease in rating compared with both other groups (*p*<0.01). However, differences in the level of devaluation were independent of deficits in goal-directed action (see *Table S15* for performance when matched for level of devaluation). There were no significant differences between groups in the average number of correct responses for the probe questions following devaluation, indicating all groups recollected the action-outcome associations.

**Figure 5.**
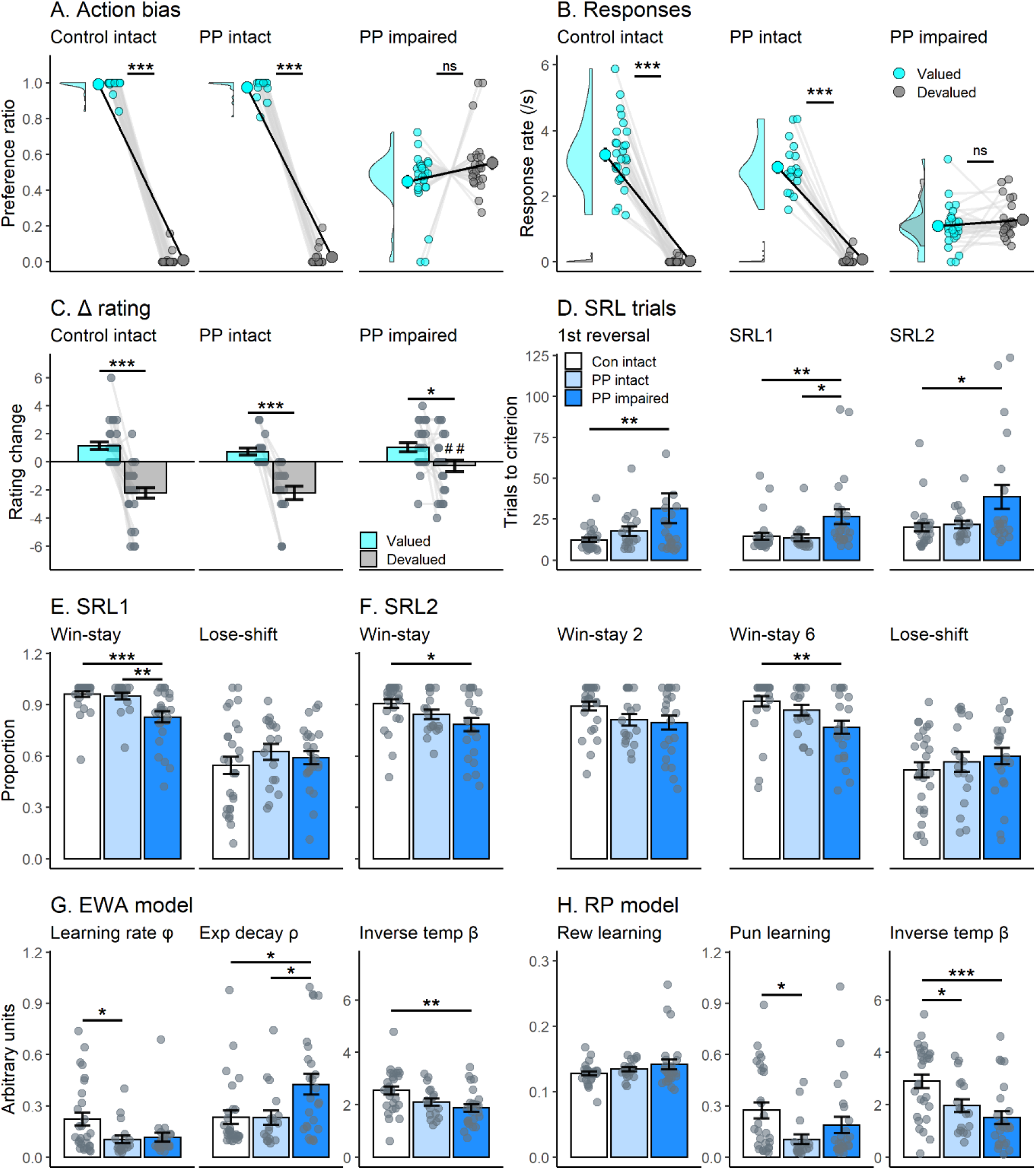
Intact and impaired goal-directed action subgroups in those with persistent psychosis. Comparison of performance in the outcome devaluation and reversal learning tasks in subjects classified as control and persistent psychosis (PP) with intact goal-directed action, and persistent psychosis with impaired goal-directed action. For outcome devaluation, persistent psychosis subjects with impaired goal-directed action did not show a preference towards the valued outcome after devaluation (**A** and **B**). Persistent psychosis subjects with impaired goal-directed action showed a significant change in their rating of the valued and devalued outcomes, highlighting that they were aware of the change in reward value (**C**). For serial reversal learning, persistent psychosis subjects with impaired goal-directed action took more trials to reach criterion (**D**) than control and persistent psychosis groups with intact goal-directed action. This was likely a consequence of a significant reduction in Win-Stay use during both the SRL1 (**E**) and SRL2 stages (**F**). Differences in computational modelling parameters were observed in both the EWA (**G**) and RP (**H**) models. Decreased learning rate (*phi*) and inverse temperature (*beta*) values were observed in both persistent psychosis groups. A significant increase in experience decay (*rho*) values in persistent psychosis subjects with impaired goal-directed action compared with both the other groups was the only cognitive process that differentiated the persistent psychosis groups. *Note: RP reward and punishment learning are inverted to match the EWA learning rate (i.e*., *1 – parameter value)*. Data are displayed as the mean ± standard error. \**p*<0.05, \*\**p*<0.01, \*\*\**p*<0.001. ^##^*p*<0.01 compared with the equivalent measure in controls.

#### 3.3.2 A decreased capacity to respond to contingency changes underlies reversal learning deficits in persistent psychosis subjects with impaired goal-directed action

General performance deficits were limited to persistent psychosis subjects with impaired goal-directed action. These were also specific to reversal learning (for all comparisons see *Table S14*), with no significant differences between groups in the trials to criterion for the initial discrimination. There were significant differences in the average trials to criterion for the first reversal (*F*_2,68_=3.3, *p*<0.05), the SRL1 stage (*F*_2,68_=6.3, *p*<0.01) and the SRL2 stage (*F*_2,61_=4.5, *p*<0.05) (**Figure 5D**). The persistent psychosis subjects with impaired goal-directed action took more trials than the control group for the first reversal (*p*<0.05) and the SRL2 stage (*p*<0.05). For the SRL1 stage, the persistent psychosis subjects with impaired goal-directed action took more trials compared with both other groups (*p*<0.05). Significant differences in the proportion of Win-stay strategy use were evident during the SRL1 (**Figure 5E**; *F*_2,68_=9.6, *p*<0.001) and SRL2 stages (**Figure 5F**; *F*_2,64_=4.1, *p*<0.05). Persistent psychosis subjects with impaired goal-directed action had a significantly lower Win-stay use on the SRL1 stage than both other groups (*p*<0.01). For the SRL2 stage, changes in Win-stay use between controls and the persistent psychosis subjects with impaired goal-directed action were greatest after winning a 6 (*p*<0.01), suggesting that higher rewards may provoke more incorrect switching.

Computational modeling highlighted that performance deficits in persistent psychosis subjects with impaired goal-directed action were associated with a unique combination of impaired processes. There were significant differences between groups for all parameters of the EWA model (**Figure 5G**). Differences in the learning rate parameter (*F*_2,68_=4.2, *p*<0.05) were driven by significant decreases in the persistent psychosis subjects with intact goal-directed action compared with the controls (*p*<0.05) and a trend towards the same in the persistent psychosis subjects with impaired goal-directed action (*p*=0.052). Differences in the experience decay parameter (*F*_2,68_=5.6, *p*<0.01) were driven by a significant increase in the persistent psychosis subjects with impaired goal-directed action compared with both other groups (*p*<0.05). Differences in the inverse temperature parameter (*F*_2,68_=5.8, *p*<0.01) were driven by a significant decrease in the persistent psychosis subjects with impaired goal-directed action compared with controls (*p*<0.01). For the RP model (**Figure 5H**), there was also a significant difference between groups for the inverse temperature (*F*_2,68_=8.5, *p*<0.001) parameter, with the control group having significantly higher inverse temperature values than both persistent psychosis groups (*p*<0.05). There was a significant difference between groups for the punishment learning parameter (*F*_2,68_=3.4, *p*<0.05), with a significant decrease in the persistent psychosis subjects with intact goal-directed action compared with the control group (*p*<0.05). Therefore, sluggish updating of experience weighting with changing contingencies (experience decay) appears to underlie the performance deficits in persistent psychosis subjects with impaired goal-directed action. In contrast, less deterministic decision-making (inverse temperature) is associated with persistent psychosis but not impairments in goal-directed action.

### 3.4 Sluggish experience updating and less deterministic choices underlie deficits in persistent psychosis subjects with impaired goal-directed action

We ran computational simulations to identify if the experience decay parameter alone could account for differences in reversal learning performance. We simulated the parameters for each group (**Figure 6**), and then using the control background, systematically altered all combinations of values from persistent psychosis subjects with impaired goal-directed action (**Figure 6** green bars). We analyzed the SRL1 stages looking at trials to criterion (**Figure 6A**) as well as Win-stay use (**Figure 6B**). A combination of increased experience decay and decreased inverse temperature was required to replicate the observed increases in trials to criterion and decreased Win-stay use. These results highlight that while persistent psychosis is associated with decreases in inverse temperature compared to controls, the additional increase in *experience decay* (specific to persistent psychosis subjects with impaired goal-directed action) is necessary to elicit performance deficits under the current task parameters.

**Figure 6.**
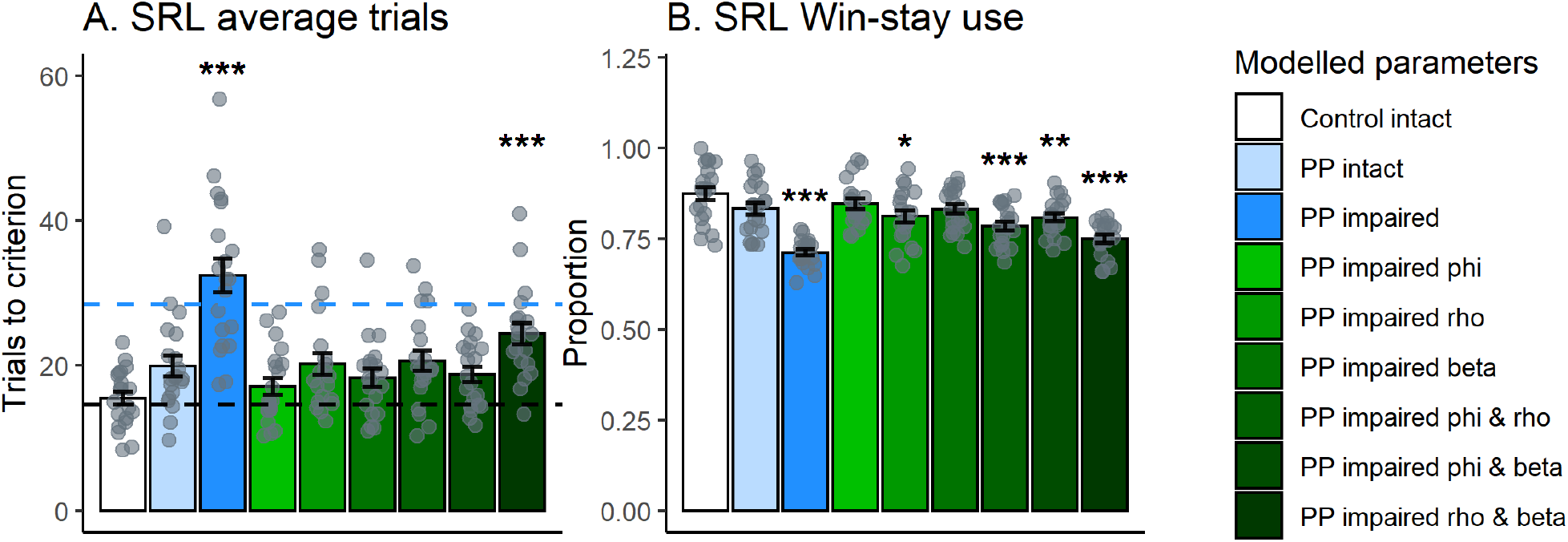
Computational simulations of decision-making performance in subgroups with intact and impaired goal-directed action. Simulations were run using the extracted parameters from the EWA model for controls with intact goal-directed action (Control intact [white]), and persistent psychosis subjects with intact (PP intact [light blue]) and impaired (PP impaired [dark blue]) goal-directed action. Compared with the control simulations, persistent psychosis subjects with impaired goal-directed action simulations took more average trials to reach criterion in the SRL stages (**A**) and used Win-stay strategies less (**B**). Simulated data reflected real performance well, demonstrated by the similar number of trials to reach criterion in the SRL1 stages (black and blue dotted lines reflect actual control intact and PP impaired averages, respectively). The green bars represent control intact parameters with one or two values substituted for PP impaired values (i.e., for light green, PP impaired *phi* with Control intact *rho* and *beta*). Only the combination of PP impaired *rho* and *beta* values (darkest green) led to both an increase in SRL trials to criterion and reduced Win-Stay use. Data are displayed as the mean ± standard error. \**p*<0.05, \*\**p*<0.01, \*\*\**p*<0.001 compared with control intact simulation (Dunnett’s test).

### 3.5 Decreases in IQ do not underly altered behavioral performance

Current IQ was lower in the persistent psychosis subjects with impaired goal-directed action compared to those with intact goal-directed action. We therefore examined persistent psychosis groups, with intact and impaired goal-directed action, matched for current and premorbid IQ (see *Table S17*). This comparison indicated that alterations in trials to criterion, Win-stay and experience decay were independent of IQ differences.

## 4. DISCUSSION

The present study extends upon existing studies (Ceaser et al., 2008; Deserno et al., 2020; Pantelis et al., 1999; Reddy et al., 2016; Waltz and Gold, 2007; Weiler et al., 2009) to show that impairments in reversal learning may be driven by a group of individuals with psychosis who feature impaired goal-directed action. Further, we observed some behavioral measures that were consistent across those with psychosis (less deterministic choices) and others specific to those with compromised goal-directed action (sluggish updating of experience weighting). Our data in those with early psychosis show that they are largely comparable to the controls but have a differing behavioral phenotype to those with persistent psychosis (e.g. changes in Lose-shift rather than Win-stay use). This suggests that either decision-making processes change, and may decline, over time in a large proportion of people with psychosis, or decision-making processes in early psychosis may indicate the subsequent trajectory of illness. Therefore, there may be avenues with which we can intervene and prevent or delay the cognitive trajectory in these individuals in early stages of psychosis.

### Decision-making in early psychosis

Those with early psychosis had relatively intact decision-making but significantly altered their responses after losses in reversal learning (i.e., decreased Lose-shifting) compared with controls. This was only observed in the SRL2 phase, where contingencies were changed from 80:20 to 80:40. We saw similar decreases in Win-stay use (6-7%) in both groups from SRL1-SRL2 highlighting that early psychosis subjects adapt similarly to controls in their response to rewarding trials. Other studies have observed that those with first-episode psychosis have a lower sensitivity to punishment than controls in reinforcement learning paradigms (Montagnese et al., 2020). This fits with our findings, in that early psychosis subjects were less likely to shift after losses (or a lack of reward in this case).

### Decision-making deficits in those with persistent psychosis

Prior studies in those with persistent psychosis have observed deficits in goal-directed action (Pantelis et al., 2004) and outcome devaluation (Morris et al., 2018; Morris et al., 2015). Like our study, no differences in the ability to understand changes in value were observed, therefore performance changes were attributed to a deficit in the ability to encode causal actions (Morris et al., 2018). In contrast to these studies (Morris et al., 2018; Morris et al., 2015), we observed intact group level outcome devaluation but the bimodal population of responding produced a weaker preference compared with control subjects. Reversal learning deficits have been observed consistently in persistent psychosis groups (Ceaser et al., 2008; Pantelis et al., 1999; Reddy et al., 2016; Waltz and Gold, 2007; Weiler et al., 2009), often accompanied by decreased Win-stay strategy use (Deserno et al., 2020; Reddy et al., 2016; Waltz et al., 2013).

### A subgroup of those with psychosis feature broad impairments in decision-making processes

We separated persistent psychosis subjects into subgroups based on intact or impaired goal-directed action. Key measures of reversal learning performance were altered in those with impaired goal-directed action. In contrast, those with persistent psychosis and intact goal-directed action performed similarly to controls in their reversal learning. Observations that certain proportions of those with persistent psychosis display impairments in reversal learning has been reported previously (Reddy et al., 2016), consistent with our study. However, the former study separated subjects based on their discrimination learning capacity, whereas we observed no differences in the trials required to complete the initial discrimination in persistent psychosis subjects with impaired goal-directed action.

We hypothesized that if the associative striatum was dysfunctional, a neurobiological mechanism thought to underlie psychosis, then we would observe impaired performance in both tasks. Our study demonstrates that impaired goal-directed action in those with persistent psychosis is accompanied by a specific reversal learning phenotype. Using computational modelling, we demonstrate that persistent psychosis subjects with impaired goal-directed action adapt to changing contingencies (i.e., reversals) slower than groups with intact goal-directed action. The EWA model was coded to reflect reversal learning-specific processes (den Ouden et al., 2013), with the experience decay parameter only impacting performance when contingencies are reversed. Differences in this parameter may also relate to striatal levels of dopamine (den Ouden et al., 2013). The increased experience decay values observed in persistent psychosis subjects with impaired goal-directed action suggests they are less willing to update their prior understanding of the associated outcome values. Similar reversal learning deficits have been observed after methylphenidate treatment (a dopamine transporter antagonist) in healthy individuals (Clatworthy et al., 2009), with those experiencing the greatest increase in dopamine in the associative striatum showing the greatest decline in reversal learning performance.

### Neurobiological mechanisms underlying impaired decision-making processes

Multiple neurobiological mechanisms could lead to this pattern of impaired performance. Corticostriatal networks may be compromised in a proportion of those with persistent psychosis and contribute to these decision-making impairments. Studies in rodents highlight a complex role for the associative striatum in decision-making, with its primary action being the maintenance and selection of optimal decision-making strategies (Ragozzino, 2007). The behavioral phenotype in persistent psychosis subjects with impaired goal-directed action suggests that increased dopamine function in the associative striatum could contribute.

However, it is commonly thought that most people with psychosis have increased associative striatal dopamine function (McCutcheon et al., 2018), yet we observe around half with impaired goal-directed action. Potentially only those with the most severe levels of dopamine/corticostriatal dysfunction feature this specific pattern of decision-making deficits.

### Psychiatric characteristics in people with persistent psychosis and intact/impaired goal-directed action

Persistent psychosis subjects had similar psychiatric characteristics overall regardless of capacity for goal-directed action, but those with impaired goal-directed action subjects had higher level of ‘difficulty in abstract thinking’ and less severe ratings of grandiosity (*Table S12*). Increased difficulty in abstract thinking makes sense given the impairments in decision-making observed in this group. How grandiosity contributes to this phenotype is unknown and clearly more work is required to understand how this relates to underlying neurobiology and functional outcomes in these individuals.

### Future directions and considerations

We recruited a modest number of those with early psychosis (N=15). However, former studies on outcome devaluation suggest this is sufficient to observe deficits in those with psychosis (Morris et al., 2018). The distinct reversal learning phenotype observed in those with early psychosis compared with persistent psychosis suggest that differing cognitive processes are impaired early after onset. This is particularly interesting in light of recent imaging work in first-episode psychosis showing structural striatal changes are dependent on antipsychotic treatment and associated with symptom reductions (Chopra et al., 2020). Future studies looking at behavioral phenotypes in early psychosis subjects that may predict subsequent declines in decision-making capacity are necessary to confirm or refute these initial findings.

## 5. CONCLUSIONS

The current study identified that a large proportion of people with persistent psychosis featured specific impairments in their decision-making capacity. Certain behavioral phenotypes appear to be consistent across those with persistent psychosis, such as a less deterministic choice strategy. Persistent psychosis subjects with impaired goal-directed action featured a decreased capacity to rapidly update their prior beliefs and associations in the face of changing contingencies. These behavioral processes are sensitive to changes in associative striatal function suggesting common neurobiology may underlie these cognitive deficits. Our observations in those with early psychosis suggest decision-making processes change after onset and decline in a proportion of those with psychosis. Thus, there may be a critical period for intervention approaches after onset in these individuals in order to maintain or delay declines in decision-making processes.

## Data Availability

All the data are available upon request.

## Contributions

Shuichi Suetani: Investigation, Writing - Original Draft, Supervision, Project administration, Funding acquisition. Andrea Baker: Conceptualization, Investigation, Resources, Data Curation, Project administration. Kelly Garner: Methodology, Software, Data Curation, Writing - Review & Editing. Peter Cosgrove: Investigation, Data Curation. Matilda Mackay-Sim: Methodology, Investigation, Writing - Review & Editing. Dan Siskind: Methodology, Writing - Review & Editing. Graham K Murray: Conceptualization, Formal analysis, Writing - Original Draft. James G Scott: Conceptualization, Resources, Writing - Original Draft, Supervision, Project administration, Funding acquisition. James P Kesby: Conceptualization, Methodology, Software, Formal analysis, Investigation, Resources, Data Curation, Writing - Original Draft, Visualization, Supervision, Project administration, Funding acquisition.

## Acknowledgements

We would like to thank all the participants, clinicians and support staff at Metro North Hospital and Health Service (MNHHS) and Metro South Addiction and Mental Health Services (MSAMHS).

## Notes

**Funding:** This work was supported by an Advance Queensland Research Fellowship (AQRF04115-16RD1 to JPK), a University of Queensland Early Career Researcher Grant (JPK), a Brisbane Diamantina Health Partners Grant (JPK and SS) and a Brain & Behavior Research Foundation Maltz Prize (JPK). AB and PC employed by, and SS and JGS are affiliated with, the Queensland Centre for Mental Health Research which receives core funding from Queensland Health. JGS is supported by a National Health and Medical Research Council Practitioner Fellowship Grant (APP1105807).

### Competing Interest Statement

The authors have declared no competing interest.

### Funding Statement

This work was supported by an Advance Queensland Research Fellowship (AQRF04115-16RD1 to JPK), a University of Queensland Early Career Researcher Grant (JPK), a Brisbane Diamantina Health Partners Grant (JPK and SS) and a Brain & Behavior Research Foundation Maltz Prize (JPK). AB and PC employed by, and SS and JGS are affiliated with, the Queensland Centre for Mental Health Research which receives core funding from Queensland Health. JGS is supported by a National Health and Medical Research Council Practitioner Fellowship Grant (APP1105807).

### Author Declarations

All procedures were approved by the Royal Brisbane and Women's Hospital, and University of Queensland Human Research Ethics Committees (HREC/17/QRBW/168).

