## Supplementary methods for "Impairments in goal-directed action and reversal learning in a proportion of individuals with psychosis: evidence for differential phenotypes in early and persistent psychosis"

**Short title:** Psychosis subgroup with select decision-making deficits

**Shuichi Suetani[1,2,3,4], Andrea Baker[1], Kelly Garner[2], Peter Cosgrove[1], Matilda Mackay-Sim[5], Dan Siskind[3], Graham K Murray[6,7,8], James G Scott[1,5,9] and James P Kesby[2,9]***

[1] Queensland Centre for Mental Health Research, Wacol, QLD 4076, Australia.
[2] Queensland Brain Institute, The University of Queensland, St Lucia, QLD 4072, Australia.
[3] Metro South Addiction and Mental Health Services, Woolloongabba, QLD 4102, Australia.
[4] School of Medicine, Griffith University, Nathan, QLD 4111, Australia.
[5] Metro North Mental Health, Royal Brisbane and Women's Hospital, Herston, QLD 4029, Australia.
[6] Department of Psychiatry, University of Cambridge, Cambridge, UK.
[7] Cambridgeshire and Peterborough NHS Foundation Trust, Cambridge, UK.
[8] Institute for Molecular Bioscience, University of Queensland, Brisbane, Australia.
[9] QIMR Berghofer Medical Research Institute, Herston, QLD 4029, Australia.

* Corresponding author:
Dr. James Kesby
Queensland Brain Institute, The University of Queensland, Brisbane, QLD 4072, Australia.

ORCID: 0000-0002-5814-8062

#### SUPPLEMENTARY METHODS

##### Procedures and experimental design

We recruited people with psychosis from Metro North Hospital and Health Services, and Metro South Addiction and Mental Health Services in Brisbane, Australia. Healthy controls were recruited from using brochures advertising the study. Participants were informed that they would receive $30 AUD for taking part in testing but could win a further $10 AUD based in the credits earned in the behavioural tasks. To avoid reduced compensation for those with psychosis and cognitive impairments, all participants were informed they had passed the 'threshold' required for the full $10 compensation. Testing order was as follows: Substance Misuse Scale, outcome devaluation instrumental training, TOPF, devaluation testing, WASI-II, reversal learning training stage, and reversal learning test stage.

##### Inclusion criteria

Those with early psychosis were recruited from Early Psychosis Clinics. Those with persistent psychosis must have been diagnosed with a Persistent Psychotic Disorder (schizophrenia, schizoaffective disorder, Bipolar I Disorder, Delusional Disorder). Participants had no organic cause pf psychosis (i.e., epilepsy, intra-cranial pathology or HIV infection), and were between the ages of 18 and 50. Subjects were classified as early psychosis if they had received less than 6 months of antipsychotic therapy.

**Serial reversal learning task**

For all stages of the reversal learning task, there were no limits on the time taken to respond. After selecting a stimulus, the outcome was presented on the screen for one second before the next trial was initiated. A running total of the 'credits' received was displayed in the bottom corner of the screen (participants were not aware of how much each credit was worth in monetary compensation). All stimulus pairs were binary images matched as closely as possible for white-black pixel ratio (see *Fig S2*), with all combinations counterbalanced.

*Initial training*

Participants were shown the following instructions on the screen: *"Two pictures will appear on the screen. On each turn, use the joystick to choose one of these pictures. The computer will tell you what credits you earned for your choice. One of the pictures will get you a reward and the other will not. The pictures will change sides randomly, so be careful to select the correct one"*. They then began a deterministic discrimination with a single reversal, whereby every correct response was rewarded (outcome of 1) and incorrect responses were not (outcome of 0). The initial discrimination contingencies were reversed after 8 consecutive correct responses were made and both stages had to be completed within 200 trials (participants got up to two attempts using unique sets of stimuli).

*Probabilistic SRL*

After successfully completing the training stage, participants were administered a probabilistic SRL task. Two instruction screens were presented, with the first restating the instruction from the training test. The second informed the participants of the probabilistic contingencies: *"Unlike before, the correct picture will not always give you a reward and sometimes the wrong picture will give you a reward. Find out which picture earns the most credits. Stick with it even if it is sometimes wrong. At some point it may change so that the other picture earns more. Only start choosing the other picture when you are sure that the rule has changed"*.

The task consisted of 11 stages, each featuring the same pair of stimuli but varying in reward rate (probabilistic) and reward value (credits awarded). These included: initial discrimination (1 stage), initial reversal (1 stage), and serial reversal learning phase 1 (SRL1; 5 stages) and serial reversal learning phase 2 (SRL2; 4 stages). For the discrimination, initial reversal and SRL1 stages, the probabilistic reward contingencies were set at 80/20, meaning that the target stimulus was rewarded 80% of the time, whereas the non-target stimulus was rewarded only 20% of the time. The reward outcomes included 1 credit for a rewarded trial and 0 credits for a non-rewarded trial. For the SRL2 stages, the probabilistic reward contingencies were set at 80/40 to increase the task difficulty, meaning that the target stimulus was rewarded 80% of the time, whereas the non-target stimulus was rewarded 40% of the time. The reward outcomes were 2 or 6 credits for a rewarded trial (of equal probability) and 0 credits for a non-rewarded trial. The addition of variable reward values was included to analyze whether the strategy a participant used was biased when receiving greater rewards on the preceding trial. Criterion for progressing to the next stage was 6 correct responses in a row. The test ended once the participant completed all 11 stages, or once 500 trials were completed. SRL1 and SRL2 trials to criterion were only included in analyses if at least 2 stages had been completed. All trials (completion of stage or not) were included in all other analyses.

*Reversal learning performance measures and strategies*

There are multiple measures of performance that can be quantified in reversal learning tasks. These include, but are not limited to, total trials to criterion, perseveration (number of errors in the first 6 trials after a reversal), and response rates (total, or for correct and incorrect responses). Other measures allow for detailed inspections of choice strategy, including whether a subject selects the same stimulus after attaining a reward (Win-stay) or whether they select the other stimulus (Win-shift). Similar strategies were calculated after losses, including whether the subject selects the same stimulus after a non-rewarded trial (Lose-stay) or selects the other stimulus (Lose-shift). These were calculated as the proportion of each strategy relative to the trials in which that strategy could be used (i.e., P.Win-stay = Total number of times the same stimulus was selected after a rewarded trial/total rewarded trials). All values were calculated for each individual stage, as well as across the SRL1 and SRL2 stages (inclusive of all combined trials).

**Serial reversal learning exclusions**

One participant (a male in the persistent psychosis group) failed to successfully complete the training stages after two attempts. Data from this participant was included in the outcome devaluation data but not for reversal learning or intact/impaired analyses.

### SUPPLEMENTARY TABLES

**Table S1.** Psychiatric characteristics and symptom assessments in those with psychosis.

| **Group** | | **Early psychosis (N = 15)** | **Persistent psychosis (N = 45)** | $F/\chi^2$ | $p$ | |
|---|---|---|---|---|---|---|
| Diagnosis (% Naff, Aff, Other) | | 7%*, 20%, | 51%, 42%, 7% | 28.5 | <0.001 | *** |
| **Medications** | | | | | | |
| Dose (chlorpromazine equivalent) | | 211.1 (171.2) | 481.0 (411.6) | 6.04 | 0.017 | * |
| Number | | 1.60 (0.91) | 4.07 (2.85) | 10.78 | 0.002 | ** |
| Anxiety (% Yes) | | 0.0% | 15.6% | 2.64 | 0.104 | |
| Mood stabilisers (% Yes) | | 26.7% | 22.2% | 0.12 | 0.724 | |
| Antidepressants (% Yes) | | 26.7% | 36.6% | 0.40 | 0.527 | |
| Substance dependence (% Yes) | | 0.0% | 11.1% | 1.82 | 0.178 | |
| **PANSS** | | | | | | |
| P1 | Delusions | 2.47 (1.36) | 2.22 (1.61) | 0.28 | 0.599 | |
| P2 | Conceptual disorganisation | 1.47 (0.99) | 1.58 (1.22) | 0.10 | 0.750 | |
| P3 | Hallucinatory Behaviour | 1.87 (1.25) | 2.51 (1.60) | 2.01 | 0.162 | |
| P4 | Excitement | 1.07 (0.26) | 1.40 (0.86) | 2.15 | 0.148 | |
| P5 | Grandiosity | 2.00 (1.36) | 2.11 (1.35) | 0.08 | 0.784 | |
| P6 | Suspiciousness/persecution | 2.33 (1.50) | 2.47 (1.49) | 0.09 | 0.765 | |
| P7 | Hostility | 1.13 (0.35) | 1.22 (0.60) | 0.29 | 0.589 | |
| **Positive Scale Total** | | **12.33 (4.67)** | **13.51 (5.69)** | **0.52** | **0.472** | |
| N1 | Blunted affect | 3.40 (1.40) | 2.87 (1.55) | 1.40 | 0.242 | |
| N2 | Emotional withdrawal | 1.93 (1.10) | 1.93 (1.25) | 0.00 | 1.000 | |
| N3 | Poor rapport | 1.27 (0.70) | 1.69 (1.10) | 1.92 | 0.171 | |
| N4 | Passive social withdrawal | 1.53 (0.83) | 1.84 (1.15) | 0.93 | 0.338 | |
| N5 | Difficulty in abstract thinking | 2.87 (1.06) | 3.51 (1.16) | 3.61 | 0.062 | |
| N6 | Lack of spontaneity/flow | 1.53 (0.99) | 1.80 (1.25) | 0.56 | 0.457 | |
| N7 | Stereotyped Thinking | 1.73 (0.88) | 1.89 (1.17) | 0.22 | 0.640 | |
| **Negative Scale Total** | | **14.27 (4.30)** | **15.53 (6.07)** | **0.56** | **0.459** | |
| G1 | Somatic concern | 2.20 (0.77) | 2.38 (1.13) | 0.32 | 0.575 | |
| G2 | Anxiety | 3.33 (1.18) | 2.64 (1.42) | 2.88 | 0.095 | |
| G3 | Guilt Feelings | 2.13 (1.46) | 1.69 (1.20) | 1.38 | 0.245 | |
| G4 | Tension | 2.47 (0.92) | 2.02 (0.94) | 2.54 | 0.116 | |
| G5 | Mannerisms and posturing | 1.47 (0.92) | 1.58 (0.99) | 0.15 | 0.703 | |
| G6 | Depression | 2.87 (0.92) | 2.42 (1.45) | 1.23 | 0.272 | |
| G7 | Motor retardation | 1.53 (0.99) | 1.56 (1.12) | 0.01 | 0.946 | |
| G8 | Uncooperativeness | 1.00 (0.00) | 1.18 (0.58) | 1.42 | 0.239 | |
| G9 | Unusual thought content | 1.60 (0.83) | 1.69 (1.00) | 0.10 | 0.757 | |
| G10 | Disorientation | 2.07 (0.96) | 2.33 (0.80) | 1.13 | 0.291 | |
| G11 | Poor attention | 1.60 (1.06) | 1.60 (1.10) | 0.00 | 1.000 | |
| G12 | Lack of judgment and insight | 3.73 (1.58) | 3.09 (1.59) | 1.85 | 0.179 | |
| G13 | Disturbance of volition | 1.40 (0.83) | 1.36 (0.74) | 0.04 | 0.846 | |
| G14 | Poor impulse control | 1.00 (0.00) | 1.18 (0.58) | 1.42 | 0.239 | |
| G15 | Preoccupation | 1.80 (1.42) | 1.84 (1.33) | 0.01 | 0.913 | |
| G16 | Active social avoidance | 1.80 (1.32) | 2.04 (1.30) | 0.40 | 0.531 | |
| **General Psychopathology Scale** | | **32.00 (8.27)** | **30.60 (7.39)** | **0.38** | **0.540** | |
| **PANSS Total** | | **58.60 (15.27)** | **59.64 (15.73)** | **0.05** | **0.823** | |

Medication classifications: Anxiety (Pregabalin, benzodiazepines), Mood stabilisers (lithium), antidepressants (selective serotonin reuptake inhibitors [SSRIs], monoamine oxidase inhibitors, Mirtazapine), and Substance dependence (Naltrexone, buprenorphine, Varenicline). *NAff*, nonaffective disorder; *Aff*, affective disorder; *AP*, antipsychotic. The data are expressed as mean (standard deviation) where applicable. *$p$<0.05, **$p$<0.01, ***$p$<0.001

**Table S2.** Demographics, IQ and substance use characteristics for control subjects aged matched to those with early psychosis.

| Group | | Controls (N = 15) | Early psychosis (N = 15) | *F*/$\chi^2$ | *p* | |
|---|---|---|---|---|---|---|
| **Demographics** | | | | | | |
| Age (years) | | 23.60 (3.29) | 21.60 (1.99) | 4.05 | 0.054 | |
| Education (years) | | 14.73 (2.58) | 12.87 (0.92) | 6.99 | 0.013 | * |
| Gender (% male) | | 46.6% | 60.0% | 0.54 | 0.464 | |
| Ethnicity (% Caucasian) | | 86.7% | 86.7% | 0.00 | 1.000 | |
| **IQ test scores** | | | | | | |
| TOPF (ss) | | 115.5 (7.5) | 115.5 (8.2) | 0.00 | 0.982 | |
| WASI-II (FISQ-2) | | 110.8 (9.6) | 105.4 (12.3) | 1.80 | 0.190 | |
| **Substance use** | | | | | | |
| Alcohol | Lifetime | 93.3% | 86.7% | 0.37 | 0.543 | |
| | 28d freq | 2.00 (1.07) | 1.40 (1.12) | 2.25 | 0.145 | |
| Cannabinoids | Lifetime | 46.7% | 66.7% | 1.22 | 0.269 | |
| | 28d freq | 0.20 (0.41) | 0.80 (1.15) | 3.64 | 0.067 | |
| Nicotine | Lifetime | 33.3% | 73.3% | 4.82 | 0.028 | * |
| | 28d freq | 0.07 (0.26) | 2.47 (2.80) | 10.93 | 0.003 | |
| Caffeine | Lifetime | 86.7% | 93.3% | 0.37 | 0.543 | |
| | 28d freq | 3.93 (2.19) | 4.00 (2.10) | 0.01 | 0.933 | |
| Amphetamines | Lifetime | 0.0% | 33.3% | 6.00 | 0.014 | * |
| | 28d freq | 0.00 (0.00) | 0.00 (0.00) | - | - | |
| Ecstasy | Lifetime | 33.3% | 46.7% | 0.56 | 0.456 | |
| | 28d freq | 0.07 (0.26) | 0.00 (0.00) | 1.00 | 0.326 | |
| Opiates | Lifetime | 0.0% | 6.7% | 1.03 | 0.309 | |
| | 28d freq | 0.00 (0.00) | 0.00 (0.00) | - | - | |
| Benzodiazepine | Lifetime | 6.7% | 20.0% | 1.15 | 0.283 | |
| | 28d freq | 0.00 (0.00) | 0.00 (0.00) | - | - | |
| Other | Lifetime | 26.7% | 40.0% | 0.60 | 0.439 | |
| | 28d freq | 0.00 (0.00) | 0.00 (0.00) | - | - | |
| Volatile | Lifetime | 0.0% | 6.7% | 1.03 | 0.309 | |
| | 28d freq | 0.00 (0.00) | 0.00 (0.00) | - | - | |

The data are expressed as mean (standard deviation) where applicable. TOPF, test of premorbid functioning; ss, standard score; WASI-II, Wechsler Abbreviated Scale of Intelligence – 2nd edition; FSIQ-2, Full-Scale IQ. *$p$<0.05.

**Table S3.** Outcome-specific devaluation in control subjects aged matched to those with early psychosis.

| Group | Controls (N = 15) | Early psychosis (N = 15) | $F/\chi^2$ | *p* |
|---|---|---|---|---|
| **Instrumental training** | | | | |
| Total trials to criterion | 6.93 (1.91) | 6.13 (0.52) | 2.46 | 0.128 |
| Total correct trials | 6.47 (1.36) | 6.07 (0.26) | 1.26 | 0.271 |
| Response rate (s) | 1.45 (0.69) | 1.38 (0.71) | 0.06 | 0.807 |
| Correct response rate (s) | 1.27 (0.67) | 1.35 (0.70) | 0.11 | 0.741 |
| **Devaluation rating changes** | | | | |
| Valued stimulus | 0.80 (1.21) [aaa] | 0.20 (1.08) [aa] | 2.05 | 0.163 |
| Devalued stimulus | -1.47 (1.36) | -1.87 (1.81) | 0.47 | 0.499 |
| Irrelevant stimulus | 0.33 (0.90) | 0.07 (1.03) | 0.57 | 0.457 |
| Motivation to earn credits | 0.13 (0.92) | 0.13 (0.92) | 0.00 | 1.000 |
| **Outcome-specific devaluation** | | | | |
| Valued response ratio | 0.76 (0.41) [a] | 0.87 (0.28) [aaa] | 0.70 | 0.410 |
| Devalued response ratio | 0.24 (0.41) | 0.13 (0.28) | 0.70 | 0.410 |
| Valued response rate (/s) | 2.55 (1.60) | 2.60 (1.52) [aa] | 0.01 | 0.944 |
| Devalued response rate | 1.04 (1.80) | 0.47 (1.11) | 1.07 | 0.309 |
| **Probe questions** | | | | |
| Average correct responses | 1.33 (0.90) | 1.73 (0.59) | 2.07 | 0.162 |

The data are expressed as mean (standard deviation) where applicable. [a]$p<0.05$, [aa]$p<0.01$, [aaa]$p<0.001$ valued outcome compared with equivalent devalued outcome (paired t-test within group).

**Table S4.** Serial reversal learning in control subjects aged matched to those with early psychosis.

| Group | Controls (N = 15) † | Early psychosis (N = 15) | $F/\chi^2$ | *p* | |
|---|---|---|---|---|---|
| **Trials to criterion** | | | | | |
| Initial discrimination | 27.33 (69.74) | 17.07 (9.80) | .032 | 0.577 | |
| First reversal | 12.33 (6.18) | 17.40 (14.24) | 1.60 | 0.217 | |
| SRL1 | 13.69 (10.77) | 16.56 (16.52) | 0.32 | 0.578 | |
| SRL2 | 18.84 (10.60) | 21.12 (17.81) | 0.17 | 0.681 | |
| **Strategy use** | | | | | |
| SRL1 Win-stay | 0.96 (0.11) | 0.93 (0.14) | 0.60 | 0.447 | |
| SRL1 Lose-shift | 0.51 (0.29) | 0.48 (0.20) | 0.10 | 0.760 | |
| SRL2 Win-stay | 0.93 (0.10) | 0.86 (0.19) | 1.30 | 0.264 | |
| SRL2 Win-stay 2 | 0.89 (0.15) | 0.85 (0.18) | 0.40 | 0.535 | |
| SRL2 Win-stay 6 | 0.96 (0.06) | 0.87 (0.20) | 2.70 | 0.112 | |
| SRL2 Lose-shift | 0.53 (0.28) | 0.35 (0.14) | 4.79 | 0.037 | * |
| **Computational modelling** | | | | | |
| EWA *phi* | 0.28 (0.22) | 0.24 (0.15) | 0.49 | 0.491 | |
| EWA *rho* | 0.22 (0.22) | 0.27 (0.29) | 0.25 | 0.622 | |
| EWA *beta* | 2.53 (0.67) | 2.06 (0.76) | 3.25 | 0.082 | |
| RP *rew* | 0.870 (0.017) | 0.864 (0.017) | 0.87 | 0.360 | |
| RP *pun* | 0.66 (0.22) | 0.71 (0.16) | 0.55 | 0.464 | |
| RP *beta* | 3.07 (1.17) | 2.13 (1.47) | 3.76 | 0.063 | |

The data are expressed as mean (standard deviation) where applicable. † for SRL2 outcomes, N = 14. *$p<0.05$.

**Table S5.** Demographics, IQ and substance use characteristics in persistent psychosis.

| **Group** | | **Controls** (N = 34) | **Persistent psychosis** (N = 45) | ***F/χ²*** | ***p*** | |
|---|---|---|---|---|---|---|
| **Demographic characteristics** | | | | | | |
| Age (years) | | 32.4 (9.9) | 31.0 (8.8) | 0.39 | 0.536 | |
| Education (years) | | 14.9 (2.7) | 11.5 (2.0) | 40.53 | <0.001 | ** |
| Gender (% male) | | 44.1% | 73.3% | 6.93 | 0.008 | ** |
| Ethnicity (% Caucasian) | | 85.29% | 66.7% | 3.55 | 0.059 | |
| **IQ test scores** | | | | | | |
| TOPF (ss) | | 112.2 (8.9) | 98.0 (14.4) | 25.43 | <0.001 | ** |
| WASI-II (FISQ-2) | | 110.1 (10.0) | 89.5 (16.9) | 40.33 | <0.001 | ** |
| **Substance use** | | | | | | |
| Alcohol | Lifetime | 97.1% | 91.1% | 1.16 | 0.282 | |
| | 28d freq | 2.26 (1.33) | 1.00 (1.26) | 18.55 | <0.001 | ** |
| Cannabinoids | Lifetime | 61.8% | 82.2% | 4.15 | 0.042 | * |
| | 28d freq | 0.32 (1.01) | 0.53 (1.27) | 0.63 | 0.431 | |
| Nicotine | Lifetime | 50.0% | 84.4% | 10.86 | <0.001 | ** |
| | 28d freq | 0.85 (2.02) | 3.87 (2.84) | 27.67 | <0.001 | ** |
| Caffeine | Lifetime | 82.4% | 95.6% | 3.71 | 0.054 | |
| | 28d freq | 4.00 (2.23) | 4.18 (2.20) | 0.13 | 0.724 | |
| Amphetamines | Lifetime | 8.8% | 53.3% | 17.06 | <0.001 | ** |
| | 28d freq | 0.00 (0.00) | 0.09 (0.47) | 1.22 | 0.272 | |
| Ecstasy | Lifetime | 23.5% | 55.6% | 8.17 | 0.004 | ** |
| | 28d freq | 0.03 (0.17) | 0.04 (0.30) | 0.69 | 0.793 | |
| Opiates | Lifetime | 2.9% | 17.8% | 4.22 | 0.040 | * |
| | 28d freq | 0.00 (0.00) | 0.00 (0.00) | - | - | |
| Benzodiazepines | Lifetime | 5.9% | 24.4% | 4.85 | 0.028 | * |
| | 28d freq | 0.00 (0.00) | 0.00 (0.00) | - | - | |
| Other | Lifetime | 26.5% | 35.6% | 0.74 | 0.390 | |
| | 28d freq | 0.06 (0.34) | 0.00 (0.00) | 0.88 | 0.418 | |
| Volatile | Lifetime | 2.9% | 17.8% | 4.22 | 0.040 | * |
| | 28d freq | 0.00 (0.00) | 0.00 (0.00) | - | - | |

The data are expressed as mean (standard deviation) where applicable. TOPF, test of premorbid functioning; ss, standard score; WASI-II, Wechsler Abbreviated Scale of Intelligence – 2nd edition; FSIQ-2, Full-Scale IQ. *$p$<0.05, **$p$<0.01, ***$p$<0.001.

**Table S6.** Outcome-specific devaluation in control subjects those with persistent psychosis.

| **Group** | **Controls** (N = 34) | **Persistent psychosis** (N = 44) | ***F/χ²*** | ***p*** | |
|---|---|---|---|---|---|
| **Instrumental training** | | | | | |
| Total trials to criterion | 6.82 (2.38) | 7.39 (2.75) | 0.90 | 0.345 | |
| Total correct trials | 6.44 (1.62) | 6.59 (1.65) | 0.16 | 0.689 | |
| Response rate (s) | 1.45 (0.63) | 2.05 (1.37) | 5.56 | 0.021 | * |
| Correct response rate (s) | 1.34 (0.63) | 1.73 (1.17) | 3.09 | 0.083 | |
| **Devaluation rating changes** | | | | | |
| Valued stimulus | 1.03 (1.45) [aaa] | 0.93 (1.39) [aaa] | 0.09 | 0.763 | |
| Devalued stimulus | -2.06 (2.03) | -1.07 (2.18) | 4.20 | 0.044 | * |
| Irrelevant stimulus | 0.00 (1.30) | -0.07 (1.81) | 0.03 | 0.853 | |
| Motivation to earn credits | 0.00 (0.98) | -0.14 (1.19) | 0.29 | 0.591 | |
| **Outcome-specific devaluation** | | | | | |
| Valued response ratio | 0.85 (0.33) [aaa] | 0.67 (0.29) [aaa] | 6.86 | 0.011 | * |
| Devalued response ratio | 0.15 (0.33) | 0.33 (0.29) | 6.86 | 0.011 | * |
| Valued response rate (/s) | 2.86 (1.42) [aaa] | 1.83 (1.12) [aaa] | 12.75 | <0.001 | ** |
| Devalued response rate | 0.59 (1.37) | 0.77 (0.73) | 0.52 | 0.472 | |
| **Probe questions** | | | | | |
| Average correct responses | 1.44 (0.79) | 1.27 (0.85) | 0.81 | 0.371 | |

The data are expressed as mean (standard deviation) where applicable. *$p$<0.05, ***$p$<0.001. [aaa]$p$<0.001 valued outcome compared with equivalent devalued outcome (paired t-test within group).

**Table S7.** Serial reversal learning in control subjects those with persistent psychosis.

| Group | Controls (N = 34) † | Persistent psychosis (N = 44) † | $F/\chi^2$ | *p* | |
|---|---|---|---|---|---|
| **Trials to criterion** | | | | | |
| Initial discrimination | 19.85 (46.53) | 18.18 (14.09) | 0.05 | 0.822 | |
| First reversal | 12.71 (6.95) | 25.45 (35.11) | 4.34 | 0.041 | * |
| SRL1 | 14.06 (9.65) | 20.87 (18.59) | 3.76 | 0.056 | |
| SRL2 | 22.64 (19.11) | 30.45 (27.35) | 1.86 | 0.177 | |
| **Strategy use** | | | | | |
| SRL1 Win-stay | 0.97 (0.08) | 0.88 (0.15) | 9.00 | 0.004 | ** |
| SRL1 Lose-shift | 0.55 (0.26) | 0.59 (0.20) | 0.80 | 0.374 | |
| SRL2 Win-stay | 0.90 (0.14) | 0.82 (0.16) | 5.39 | 0.023 | * |
| SRL2 Win-stay 2 | 0.88 (0.16) | 0.81 (0.18) | 3.49 | 0.066 | |
| SRL2 Win-stay 6 | 0.92 (0.16) | 0.82 (0.17) | 6.22 | 0.015 | * |
| SRL2 Lose-shift | 0.52 (0.25) | 0.57 (0.25) | 0.96 | 0.330 | |
| **Computational modelling** | | | | | |
| EWA *phi* | 0.22 (0.19) | 0.12 (0.13) | 7.19 | 0.009 | ** |
| EWA *rho* | 0.22 (0.19) | 0.34 (0.27) | 4.57 | 0.036 | * |
| EWA *beta* | 2.53 (0.78) | 2.02 (0.73) | 8.56 | 0.005 | ** |
| RP *rew* | 0.87 (0.02) | 0.86 (0.03) | 4.74 | 0.033 | * |
| RP *pun* | 0.72 (0.23) | 0.84 (0.21) | 4.96 | 0.029 | * |
| RP *beta* | 2.90 (1.35) | 1.80 (1.34) | 12.75 | <0.001 | ** |

The data are expressed as mean (standard deviation) where applicable. **†** for SRL2 outcomes, N = 33 for control and N = 41 for persistent psychosis groups. *$p<0.05$, **$p<0.01$, ***$p<0.001$.

**Table S8.** Demographics, IQ and substance use characteristics for control subjects split for intact or impaired goal-directed action.

| **Group** | | **Control** | **Control** |
|---|---|---|---|
| **Goal-directed action** | | **intact (N = 28)** | **impaired (N = 6)** |
| **Demographics** | | | |
| Age (years) | | 32.3 (10.0) | 32.5 (10.4) |
| Education (years) | | 14.7 (2.8) | 15.5 (2.0) |
| Gender (% male) | | 35.7% | 83.3% |
| Ethnicity (% Caucasian) | | 89.3% | 66.7% |
| **IQ test scores** | | | |
| TOPF (ss) | | 111.5 (9.4) | 115.2 (6.4) |
| WASI-II (FISQ-2) | | 110.1 (10.7) | 110.3 (5.9) |
| **Substance use** | | | |
| Alcohol | Lifetime | 100.0% | 83.3% |
| | 28d freq | 2.25 (1.29) | 2.33 (1.63) |
| Cannabinoids | Lifetime | 64.3% | 50.0% |
| | 28d freq | 0.21 (0.63) | 0.83 (2.04) |
| Nicotine | Lifetime | 50.0% | 50.0% |
| | 28d freq | 0.82 (1.96) | 1.00 (2.45) |
| Caffeine | Lifetime | 82.1% | 83.3% |
| | 28d freq | 3.93 (2.26) | 4.33 (2.25) |
| Amphetamines | Lifetime | 7.1% | 16.7% |
| | 28d freq | — | — |
| Ecstasy | Lifetime | 21.4% | 33.3% |
| | 28d freq | 0.04 (0.19) | — |
| Opiates | Lifetime | 3.6% | 0.0% |
| | 28d freq | — | — |
| Benzodiazepine | Lifetime | 7.1% | 0.0% |
| | 28d freq | — | — |
| Other | Lifetime | 25.0% | 33.3% |
| | 28d freq | 0.07 (0.38) | — |
| Volatile | Lifetime | 0.0% | 16.7% |
| | 28d freq | — | — |

The data are expressed as mean (standard deviation) where applicable. TOPF, test of premorbid functioning; ss, standard score; WASI-II, Wechsler Abbreviated Scale of Intelligence – 2$^{nd}$ edition; FSIQ-2, Full-Scale IQ.

**Table S9.** Demographics, IQ and substance use characteristics for those with early psychosis split for intact or impaired goal-directed action.

| Group<br>Goal-directed action | | Early psychosis<br>intact<br>(N = 12) | Early psychosis<br>impaired<br>(N = 3) |
|---|---|---|---|
| **Demographics** | | | |
| Age (years) | | 21.5 (2.2) | 22.0 (1.0) |
| Education (years) | | 12.8 (1.0) | 13.3 (0.6) |
| Gender (% male) | | 50.0% | 100.0% |
| Ethnicity (% Caucasian) | | 83.3% | 100.0% |
| **IQ test scores** | | | |
| TOPF (ss) | | 117.4 (8.0) | 108.0 (2.0) |
| WASI-II (FISQ-2) | | 107.9 (11.0) | 95.3 (14.0) |
| **Substance use** | | | |
| Alcohol | Lifetime | 83.3% | 100.0% |
| | 28d freq | 1.17 (1.03) | 2.33 (1.15) |
| Cannabinoids | Lifetime | 58.3% | 100.0% |
| | 28d freq | 0.92 (1.24) | 0.33 (0.58) |
| Nicotine | Lifetime | 66.7% | 100.0% |
| | 28d freq | 2.75 (2.93) | 1.33 (2.31) |
| Caffeine | Lifetime | 91.7% | 100.0% |
| | 28d freq | 3.83 (2.25) | 4.67 (1.53) |
| Amphetamines | Lifetime | 33.3% | 33.3% |
| | 28d freq | — | — |
| Ecstasy | Lifetime | 50.0% | 33.3% |
| | 28d freq | — | — |
| Opiates | Lifetime | 0.0% | 33.3% |
| | 28d freq | — | — |
| Benzodiazepine | Lifetime | 16.7% | 33.3% |
| | 28d freq | — | — |
| Other | Lifetime | 41.7% | 33.3% |
| | 28d freq | — | — |
| Volatile | Lifetime | 8.3% | 0.0% |
| | 28d freq | — | — |

The data are expressed as mean (standard deviation) where applicable. TOPF, test of premorbid functioning; ss, standard score; WASI-II, Wechsler Abbreviated Scale of Intelligence – 2nd edition; FSIQ-2, Full-Scale IQ.

**Table S10.** Psychiatric characteristics and symptom assessments for those with early psychosis split for intact or impaired goal-directed action.

| **Group** | **Early psychosis** | **Early psychosis** |
|---|---|---|
| **Goal-directed action** | **intact (N = 12)** | **impaired (N = 3)** |
| **Medications** | | |
| Dose (CPZ equivalent) | 202.7 (178.2) | 245.0 (168.5) |
| Number | 1.67 (0.98) | 1.33 (0.58) |
| Anxiety (% Yes) | 0.0% | 0.0% |
| Mood stabilisers (% Yes) | 25.0% | 33.3% |
| Antidepressants (% Yes) | 33.3% | 0.0% |
| Dependence (% Yes) | 0.0% | 0.0% |
| **PANSS** | | |
| Positive Scale Total | 11.92 (5.16) | 14.00 (1.00) |
| Negative Scale Total | 14.42 (4.40) | 13.67 (4.73) |
| Psychopathology Total | 33.00 (8.84) | 28.00 (4.36) |
| PANSS Total | 59.33 (16.74) | 55.67 (8.74) |

Medication classifications: Anxiety (Pregabalin, benzodiazepines), Mood stabilisers (lithium), antidepressants (selective serotonin reuptake inhibitors [SSRIs], monoamine oxidase inhibitors, Mirtazapine), and Substance dependence (Naltrexone, buprenorphine, Varenicline). CPZ, chlorpromazine. The data are expressed as mean (standard deviation) where applicable.

**Table S11.** Demographics, IQ and substance use characteristics for those with persistent psychosis split for intact or impaired goal-directed action.

| **Group** | | **Control** | **Persistent psychosis** | **Persistent psychosis** | $F/\chi^2$ | $p$ |
|---|---|---|---|---|---|---|
| **Goal-directed action** | | **intact (N = 28)** | **intact (N = 18)** | **impaired (N = 25)** | | |
| **Demographic characteristics** | | | | | | |
| Age (years) | | 32.3 (10.0) | 31.6 (9.6) | 31.2 (8.4) | 0.10 | 0.902 |
| Education (years) | | 14.7 (2.8) | 12.4 (1.5) ## | 10.9 (2.1) ### | 18.75 | <0.001 |
| Gender (% male) | | 35.7% | 77.8% | 68.0% | 9.61 | 0.008 |
| Ethnicity (% Caucasian) | | 89.3% | 83.3% | 56.0% | 8.74 | 0.013 |
| **IQ test scores** | | | | | | |
| TOPF (ss) | | 111.5 (9.4) | 103.8 (9.8) | 94.5 (15.1) ### | 13.81 | <0.001 |
| WASI-II (FISQ-2) | | 110.1 (10.7) | 98.8 (14.9) # | 83.5 (14.5) ** | 26.62 | <0.001 |
| **Substance use** | | | | | | |
| Alcohol | Lifetime | 100.0% | 88.9% | 96.0% | 3.35 | 0.188 |
| | 28d freq | 2.25 (1.29) | 1.33 (1.46) | 0.84 (1.11) ### | 8.33 | 0.001 |
| Cannabinoids | Lifetime | 64.3% | 72.2% | 88.0% | 4.00 | 0.135 |
| | 28d freq | 0.21 (0.63) | 0.67 (1.57) | 0.48 (1.08) | 1.00 | 0.375 |
| Nicotine | Lifetime | 50.0% | 83.3% | 84.0% # | 9.26 | 0.010 |
| | 28d freq | 0.82 (1.96) | 3.33 (2.91) ## | 4.08 (2.86) ### | 11.72 | <0.001 |
| Caffeine | Lifetime | 82.1% | 94.4% | 96.0% | 3.36 | 0.187 |
| | 28d freq | 3.93 (2.26) | 4.17 (2.28) | 4.04 (2.21) | 0.06 | 0.940 |
| Amphetamines | Lifetime | 7.1% | 44.4% # | 60.0% # | 17.08 | <0.001 |
| | 28d freq | 0.00 (0.00) | 0.00 (0.00) | 0.16 (0.62) | 1.51 | 0.229 |
| Ecstasy | Lifetime | 21.4% | 61.1% # | 52.0% | 8.57 | 0.014 |
| | 28d freq | 0.04 (0.19) | 0.11 (0.47) | 0.00 (0.00) | 0.94 | 0.395 |
| Opiates | Lifetime | 3.6% | 16.7% | 20.0% | 3.57 | 0.168 |
| | 28d freq | 0.00 (0.00) | 0.00 (0.00) | 0.00 (0.00) | - | - |
| Benzodiazepines | Lifetime | 7.1% | 38.9% # | 16.0% | 7.52 | 0.023 |
| | 28d freq | 0.00 (0.00) | 0.00 (0.00) | 0.00 (0.00) | - | - |
| Other | Lifetime | 25.0% | 44.4% | 28.0% | 2.10 | 0.350 |
| | 28d freq | 0.07 (0.38) | 0.00 (0.00) | 0.00 (0.00) | 0.76 | 0.470 |
| Volatile | Lifetime | 0.0% | 22.2% # | 16.0% | 6.276 | 0.043 |
| | 28d freq | 0.00 (0.00) | 0.00 (0.00) | 0.00 (0.00) | - | - |

The data are expressed as mean (standard deviation) where applicable. TOPF, test of premorbid functioning; ss, standard score; WASI-II, Wechsler Abbreviated Scale of Intelligence – 2nd edition; FSIQ-2, Full-Scale IQ. **$p$<0.01 compared with all groups; #$p$<0.05, ##$p$<0.01, ###$p$<0.001 compared with Controls.

**Table S12.** Psychiatric characteristics and symptom assessments for those with persistent psychosis split for intact or impaired goal-directed action.

| **Group**<br><br>**Goal-directed action** | | **Persistent psychosis**<br>**intact**<br>**(N = 18)** | **Persistent psychosis**<br>**impaired**<br>**(N = 25)** | ***F/*$\chi^2$** | ***p*** | |
|---|---|---|---|---|---|---|
| Diagnosis (% Naff, Aff, Other) | | 56%, 44%, 0% | 44%, 39%, 17% | 4.51 | 0.105 | |
| **Medications** | | | | | | |
| Dose (chlorpromazine equivalent) | | 424.8 (225.9) | 521.8 (513.8) | 0.56 | 0.458 | |
| Number | | 3.61 (2.97) | 4.48 (2.80) | 0.96 | 0.334 | |
| Anxiety (% Yes) | | 11.1% | 20.0% | 0.61 | 0.436 | |
| Mood stabilisers (% Yes) | | 27.8% | 16.0% | 0.88 | 0.349 | |
| Antidepressants (% Yes) | | 33.3% | 40.0% | 0.20 | 0.655 | |
| Substance dependence (% Yes) | | 16.7% | 8.0% | 0.77 | 0.382 | |
| **PANSS** | | | | | | |
| P1 | Delusions | 2.39 (1.72) | 2.20 (1.58) | 0.14 | 0.711 | |
| P2 | Conceptual disorganisation | 1.72 (1.41) | 1.52 (1.12) | 0.28 | 0.603 | |
| P3 | Hallucinatory Behaviour | 2.44 (1.85) | 2.60 (1.47) | 0.09 | 0.761 | |
| P4 | Excitement | 1.61 (1.04) | 1.28 (0.74) | 1.50 | 0.227 | |
| P5 | Grandiosity | 2.72 (1.53) | 1.64 (1.08) | 7.46 | 0.009 | ** |
| P6 | Suspiciousness/persecution | 2.39 (1.58) | 2.56 (1.50) | 0.13 | 0.720 | |
| P7 | Hostility | 1.06 (0.24) | 1.36 (0.76) | 2.71 | 0.108 | |
| **Positive Scale Total** | | **14.33 (6.13)** | **13.16 (5.59)** | **0.43** | **0.518** | |
| N1 | Blunted affect | 2.56 (1.62) | 3.08 (1.44) | 1.25 | 0.270 | |
| N2 | Emotional withdrawal | 1.67 (1.14) | 2.08 (1.29) | 1.19 | 0.283 | |
| N3 | Poor rapport | 1.39 (1.04) | 1.84 (1.07) | 1.91 | 0.174 | |
| N4 | Passive social withdrawal | 1.89 (1.28) | 1.88 (1.09) | 0.00 | 0.981 | |
| N5 | Difficulty in abstract thinking | 2.94 (1.00) | 3.88 (1.01) | 9.03 | 0.005 | ** |
| N6 | Lack of spontaneity/flow | 1.39 (1.14) | 2.00 (1.15) | 2.95 | 0.093 | |
| N7 | Stereotyped Thinking | 1.94 (1.30) | 1.72 (1.02) | 0.40 | 0.530 | |
| **Negative Scale Total** | | **13.78 (6.30)** | **16.48 (5.61)** | **2.19** | **0.146** | |
| G1 | Somatic concern | 2.61 (1.20) | 2.24 (1.09) | 1.12 | 0.296 | |
| G2 | Anxiety | 2.56 (1.42) | 2.84 (1.40) | 0.42 | 0.518 | |
| G3 | Guilt Feelings | 1.83 (1.38) | 1.56 (1.08) | 0.53 | 0.471 | |
| G4 | Tension | 1.89 (0.96) | 2.20 (0.91) | 1.16 | 0.288 | |
| G5 | Mannerisms and posturing | 1.50 (0.86) | 1.64 (1.11) | 0.20 | 0.658 | |
| G6 | Depression | 2.56 (1.54) | 2.44 (1.42) | 0.07 | 0.801 | |
| G7 | Motor retardation | 1.33 (1.03) | 1.60 (1.00) | 0.73 | 0.399 | |
| G8 | Uncooperativeness | 1.00 (0.00) | 1.24 (0.66) | 2.34 | 0.134 | |
| G9 | Unusual thought content | 1.89 (1.13) | 1.60 (0.91) | 0.86 | 0.360 | |
| G10 | Disorientation | 2.17 (0.86) | 2.52 (0.71) | 2.17 | 0.149 | |
| G11 | Poor attention | 1.39 (1.24) | 1.80 (1.00) | 1.44 | 0.237 | |
| G12 | Lack of judgment and insight | 2.78 (1.59) | 3.24 (1.64) | 0.85 | 0.362 | |
| G13 | Disturbance of volition | 1.50 (0.92) | 1.28 (0.61) | 0.88 | 0.353 | |
| G14 | Poor impulse control | 1.22 (0.65) | 1.16 (0.55) | 0.12 | 0.736 | |
| G15 | Preoccupation | 1.72 (1.36) | 1.84 (1.21) | 0.09 | 0.767 | |
| G16 | Active social avoidance | 1.83 (1.42) | 2.20 (1.22) | 0.82 | 0.371 | |
| **General Psychopathology Scale** | | **29.78 (9.72)** | **31.40 (5.53)** | **0.48** | **0.491** | |
| **PANSS Total** | | **57.89 (19.70)** | **61.04 (12.81)** | **0.40** | **0.528** | |

Medication classifications: Anxiety (Pregabalin, benzodiazepines), Mood stabilisers (lithium), antidepressants (selective serotonin reuptake inhibitors [SSRIs], monoamine oxidase inhibitors, Mirtazapine), and Substance dependence (Naltrexone, buprenorphine, Varenicline). NAff, nonaffective disorder; Aff, affective disorder. The data are expressed as mean (standard deviation) where applicable. $^{**}p<0.01$.

**Table S13.** Outcome-specific devaluation in controls and those with persistent psychosis split for intact or impaired goal-directed action.

| **Group** | **Control** | **Persistent psychosis** | **Persistent psychosis** | ***F*/$\chi^2$** | ***p*** |
|---|---|---|---|---|---|
| **Goal-directed action** | **intact (N = 28)** | **intact (N = 18)** | **impaired (N = 25)** | | |
| **Instrumental training** | | | | | |
| Total trials to criterion | 6.21 (0.63) | 7.00 (2.87) | 7.44 (2.50) | 2.31 | 0.107 |
| Total correct trials | 6.07 (0.38) | 6.33 (1.41) | 6.68 (1.77) | 1.47 | 0.237 |
| Response rate (s) | 1.48 (0.64) | 2.03 (1.15) | 1.80 (0.86) | 2.38 | 0.100 |
| Correct response rate (s) | 1.38 (0.67) | 1.89 (1.20) | 1.44 (0.65) | 2.27 | 0.111 |
| **Devaluation rating changes** | | | | | |
| Valued stimulus | 1.14 (1.51) [aaa] | 0.72 (1.02) [aaa] | 1.04 (1.62) [a] | 0.48 | 0.622 |
| Devalued stimulus | -2.21 (1.93) | -2.22 (1.99) | -0.28 (2.01) ** | 7.80 | 0.001 |
| Irrelevant stimulus | 0.04 (1.43) | -0.28 (2.24) | 0.08 (1.50) | 0.27 | 0.766 |
| Motivation to earn credits | 0.14 (0.89) | 0.28 (0.75) | -0.48 (1.36) | 3.43 | 0.038 |
| **Outcome-specific devaluation** | | | | | |
| Valued response ratio | 0.99 (0.03) [aaa] | 0.98 (0.05) [aaa] | 0.45 (0.17) *** | 47.05 | <0.001 |
| Devalued response ratio | 0.01 (0.03) | 0.02 (0.05) | 0.55 (0.17) *** | 112.91 | <0.001 |
| Valued response rate (/s) | 3.26 (1.03) [aaa] | 2.88 (0.77) [aaa] | 1.08 (0.66) *** | 203.38 | <0.001 |
| Devalued response rate | 0.02 (0.06) | 0.07 (0.16) | 1.27 (0.54) *** | 203.38 | <0.001 |
| **Probe questions** | | | | | |
| Average correct responses | 1.61 (0.69) | 1.56 (0.51) | 1.12 (0.97) | 3.05 | 0.054 |

The data are expressed as mean (standard deviation) where applicable. ***p*<0.01, ****p*<0.001 compared with all groups; [#]*p*<0.05, [##]*p*<0.01, [###]*p*<0.001 compared with Controls; [a]*p*<0.05, [aaa]*p*<0.001 valued outcome compared with equivalent devalued outcome (paired t-test within group).

**Table S14.** Serial reversal learning in controls and those with persistent psychosis split for intact or impaired goal-directed action.

| **Group** | **Control** | **Persistent psychosis** | **Persistent psychosis** | ***F/χ²*** | ***p*** |
|---|---|---|---|---|---|
| **Goal-directed action** | **intact (N = 28) †** | **intact (N = 18)** | **impaired (N = 25) †** | | |
| **Trials to criterion** | | | | | |
| Initial discrimination | 21.89 (51.16) | 17.22 (13.74) | 19.12 (14.79) | 0.11 | 0.897 |
| First reversal | 12.29 (6.83) | 17.78 (11.89) | 31.60 (44.89) # | 3.34 | 0.042 |
| SRL1 | 14.59 (10.48) | 13.57 (8.21) | 26.51 (22.49) * | 5.12 | 0.009 |
| SRL2 | 20.08 (13.33) | 21.74 (9.96) | 38.60 (35.54) # | 4.46 | 0.016 |
| **Strategy use** | | | | | |
| SRL1 Win-stay | 0.96 (0.09) | 0.95 (0.09) | 0.83 (0.16) ** | 9.57 | <0.001 |
| SRL1 Lose-shift | 0.55 (0.26) | 0.62 (0.20) | 0.59 (0.18) | 0.73 | 0.484 |
| SRL2 Win-stay | 0.91 (0.13) | 0.84 (0.12) | 0.78 (0.19) # | 4.07 | 0.022 |
| SRL2 Win-stay 2 | 0.89 (0.14) | 0.81 (0.15) | 0.80 (0.20) | 2.47 | 0.092 |
| SRL2 Win-stay 6 | 0.92 (0.17) | 0.87 (0.13) | 0.77 (0.19) # | 5.02 | 0.009 |
| SRL2 Lose-shift | 0.52 (0.23) | 0.57 (0.25) | 0.60 (0.23) | 0.73 | 0.485 |
| **Computational modelling** | | | | | |
| EWA *phi* | 0.22 (0.21) | 0.10 (0.10) # | 0.12 (0.13) | 4.18 | 0.019 |
| EWA *rho* | 0.23 (0.20) | 0.23 (0.17) | 0.43 (0.29) ** | 5.55 | 0.006 |
| EWA *beta* | 2.54 (0.81) | 2.10 (0.59) | 1.88 (0.70) ## | 5.80 | 0.005 |
| RP *rew* | 0.87 (0.02) | 0.87 (0.01) | 0.86 (0.04) | 1.95 | 0.151 |
| RP *pun* | 0.73 (0.25) | 0.90 (0.12) # | 0.81 (0.23) | 3.40 | 0.039 |
| RP *beta* | 2.91 (1.38) | 1.97 (1.06) # | 1.51 (1.24) ## | 8.50 | <0.001 |

The data are expressed as mean (standard deviation) where applicable. † for SRL2 outcomes, N = 27 for control intact and N = 22 for persistent psychosis impaired groups. *$p<0.05$, **$p<0.01$ compared with all groups; #$p<0.05$, ##$p<0.01$ compared with Controls.

**Table S15.** Outcome-specific devaluation in a subgroup of control subjects **matched for rating changes towards the devalued stimuli** with persistent psychosis subjects with impaired goal-directed action.

| **Group** | **Control** | **Persistent psychosis** | ***F/χ²*** | ***p*** | |
|---|---|---|---|---|---|
| **Goal-directed action** | **intact (N = 15)** | **impaired (N = 25)** | | | |
| **Instrumental training** | | | | | |
| Total trials to criterion | 6.20 (0.77) | 7.44 (2.50) | 3.46 | 0.071 | |
| Total correct trials | 6.13 (0.52) | 6.68 (1.77) | 1.35 | 0.253 | |
| Response rate (s) | 1.40 (0.67) | 1.80 (0.86) | 2.44 | 0.127 | |
| Correct response rate (s) | 1.39 (0.67) | 1.44 (0.65) | 0.04 | 0.840 | |
| **Devaluation rating changes** | | | | | |
| Valued stimulus | 1.20 (1.70) | 1.04 (1.62) | 0.09 | 0.768 | |
| ***Devalued stimulus*** | ***-0.93 (1.03)*** | ***-0.28 (2.01)*** | ***1.36*** | ***0.251*** | |
| Irrelevant stimulus | 0.27 (0.88) | 0.08 (1.50) | 0.19 | 0.664 | |
| Motivation to earn credits | 0.40 (1.12) | -0.48 (1.36) | 4.46 | 0.041 | * |
| **Outcome-specific devaluation** | | | | | |
| Valued response ratio | 0.99 (0.04) [aaa] | 0.45 (0.17) [aaa] | 142.46 | <0.001 | ** |
| Devalued response ratio | 0.01 (0.04) | 0.55 (0.17) | 142.46 | <0.001 | ** |
| Valued response rate (/s) | 2.88 (0.73) [a] | 1.08 (0.66) | 63.47 | <0.001 | ** |
| Devalued response rate | 0.02 (0.07) | 1.27 (0.54) | 79.83 | <0.001 | ** |
| **Probe questions** | | | | | |
| Average correct responses | 1.67 (0.62) | 1.12 (0.97) | 3.81 | 0.058 | |

The data are expressed as mean (standard deviation) where applicable.
*$p$<0.05, ***$p$<0.001. [a]$p$<0.05, [aaa]$p$<0.001 valued outcome compared with equivalent devalued outcome (paired t-test within group).

**Table S16.** Serial reversal learning in a sub cohort of control intact subjects **matched for rating changes towards the devalued stimuli** with PP impaired subjects (*see Table S15*).

| **Group** | **Control** | **Persistent psychosis** | $F/\chi^2$ | ***p*** | |
|---|---|---|---|---|---|
| **Goal-directed action** | **intact (N = 15) †** | **impaired (N = 24) †** | | | |
| **Trials to criterion** | | | | | |
| Initial discrimination | 28.87 (69.66) | 19.12 (14.79) | 0.51 | 0.479 | |
| First reversal | 12.27 (8.31) | 31.60 (44.89) | 2.17 | 0.149 | |
| SRL1 | 16.37 (14.11) | 26.51 (22.49) | 2.44 | 0.127 | |
| SRL2 | 19.27 (10.17) | 38.60 (35.54) | 3.89 | 0.057 | |
| **Strategy use** | | | | | |
| SRL1 Win-stay | 0.95 (0.11) | 0.83 (0.16) | 6.17 | 0.018 | * |
| SRL1 Lose-shift | 0.51 (0.25) | 0.59 (0.18) | 1.54 | 0.223 | |
| SRL2 Win-stay | 0.91 (0.12) | 0.78 (0.19) | 3.54 | 0.069 | |
| SRL2 Win-stay 2 | 0.88 (0.13) | 0.80 (0.20) | 0.91 | 0.349 | |
| SRL2 Win-stay 6 | 0.95 (0.12) | 0.77 (0.19) | 7.40 | 0.010 | * |
| SRL2 Lose-shift | 0.52 (0.23) | 0.60 (0.23) | 0.85 | 0.363 | |
| **Computational modelling** | | | | | |
| EWA *phi* | 0.24 (0.23) | 0.12 (0.13) | 4.74 | 0.036 | * |
| EWA *rho* | 0.24 (0.23) | 0.43 (0.29) | 4.25 | 0.046 | * |
| EWA *beta* | 2.55 (0.79) | 1.88 (0.70) | 7.95 | 0.008 | ** |
| RP *rew* | 0.72 (0.23) | 0.81 (0.23) | 1.46 | 0.235 | |
| RP *pun* | 0.87 (0.02) | 0.86 (0.04) | 1.45 | 0.236 | |
| RP *beta* | 3.00 (1.45) | 1.51 (1.24) | 11.90 | <0.001 | ** |

The data are expressed as mean (standard deviation) where applicable. **†** for SRL2 outcomes, N = 14 for control intact and N = 20 for persistent psychosis impaired groups. *$p$<0.05, **$p$<0.01, ***$p$<0.001.

**Table S17.** Behavioural differences in those with persistent psychosis split for intact or impaired goal-directed action and **matched for IQ**.

| **Group**<br><br>**Goal-directed action** | **Persistent psychosis intact (N = 17)** | **Persistent psychosis impaired (N = 17)** | ***F*** | ***p*** | |
|---|---|---|---|---|---|
| **General characteristics** | | | | | |
| Age (years) | 32.2 (9.5) | 31.2 (9.9) | 0.10 | 0.753 | |
| Dose (CPZ equivalent) | 449.8 (205.6) | 575.6 (608.6) | 0.65 | 0.425 | |
| **IQ test scores** | | | | | |
| TOPF (ss) | 103.3 (9.8) | 97.3 (14.8) | 1.95 | 0.172 | |
| WASI-II (FISQ-2) | 96.6 (11.8) | 91.5 (9.6) | 1.89 | 0.179 | |
| **Outcome-specific** | | | | | |
| Valued response ratio | 0.97 (0.05) | 0.49 (0.15) | 160.27 | <0.001 | *** |
| Valued response rate (/s) | 2.93 (0.76) | 1.24 (0.7) | 44.62 | <0.001 | *** |
| **Serial reversal learning** | | | | | |
| SRL1 (trials to criterion) | 13.88 (8.35) | 26.13 (20.46) | 5.22 | 0.029 | * |
| SRL1 Win-stay | 0.95 (0.09) | 0.83 (0.16) | 7.77 | 0.009 | ** |
| SRL1 Lose-shift | 0.63 (0.2) | 0.63 (0.16) | 0.00 | 0.991 | |
| **Computational modelling** | | | | | |
| EWA phi | 0.16 (0.17) | 0.09 (0.06) | 0.55 | 0.463 | |
| EWA rho | 0.38 (0.21) | 0.48 (0.28) | 4.59 | 0.040 | * |
| EWA beta | 2.07 (0.56) | 1.96 (0.71) | 1.87 | 0.181 | |

The data are expressed as mean (standard deviation) where applicable. CPZ, chlorpromazine; TOPF, test of premorbid functioning; ss, standard score; WASI-II, Wechsler Abbreviated Scale of Intelligence – $2^{nd}$ edition; FSIQ-2, Full-Scale IQ. **p*<0.05, ***p*<0.01, ****p*<0.001.

#### SUPPLEMENTARY FIGURES

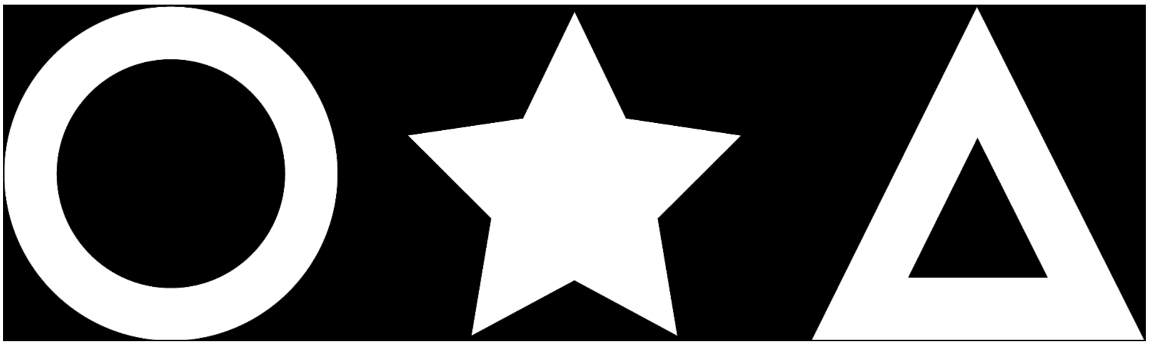

**Figure S1. Stimuli used for outcome-specific devaluation tokens.** All stimuli were matched for white:black pixel ratio.

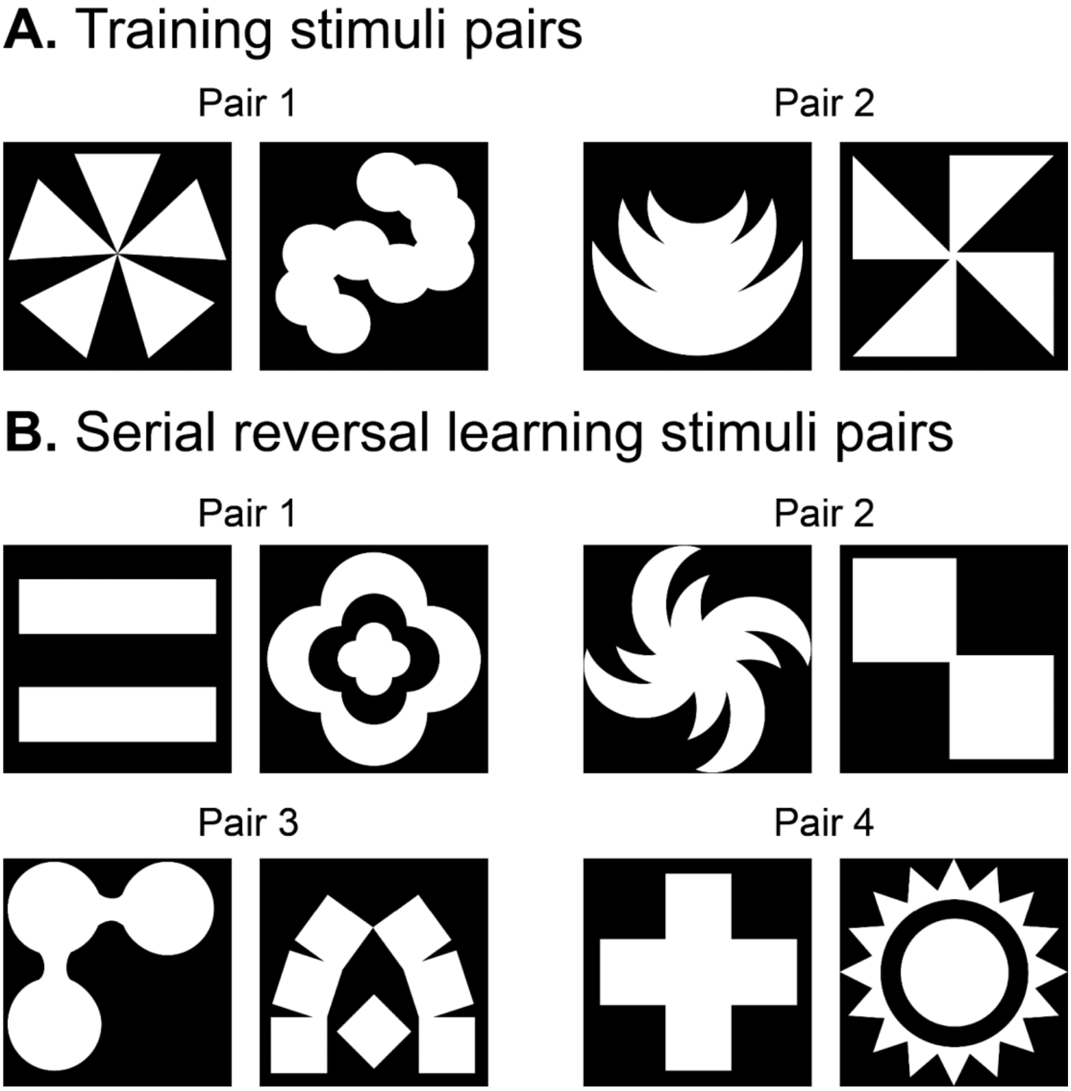


**Figure S2. Visual stimuli pairs used for serial reversal learning.** Two pairs of stimuli were used for the training stage (**A**). Four pairs of stimuli were used for the serial reversal learning test stage (**B**). Pair sets and stimuli assigned as the initial 'correct' stimuli were counterbalanced between groups. All stimuli were matched for white:black pixel ratio.

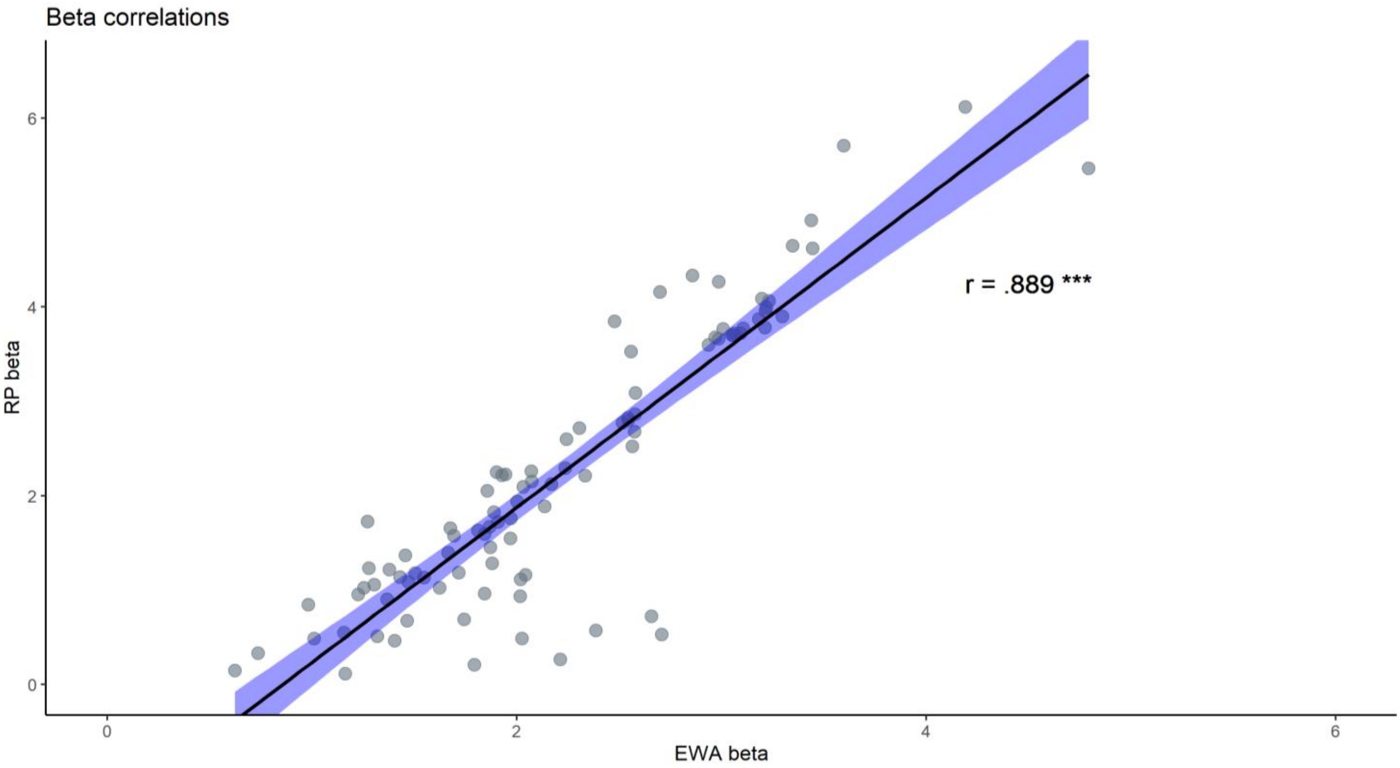


**Figure S3. Correlation between beta values for the experience weighted attraction (EWA) and reinforcement learning (RP) models.** The beta values (inverse temperature) estimated by both models correlated highly ($r_{(94)} = 0.89$, $p < 0.001$), supporting that both models accurately estimate underlying cognitive processes. Standard error is represented by the blue shading.
